# Data-driven hypothesis generation among inexperienced clinical researchers: A comparison of secondary data analyses with visualization (VIADS) and other tools

**DOI:** 10.1101/2023.05.30.23290719

**Authors:** Xia Jing, James J. Cimino, Vimla L. Patel, Yuchun Zhou, Jay H. Shubrook, Sonsoles De Lacalle, Brooke N. Draghi, Mytchell A. Ernst, Aneesa Weaver, Shriram Sekar, Chang Liu

## Abstract

**Objectives:** To compare how clinical researchers generate data-driven hypotheses with a visual interactive analytic tool (VIADS, a <u>v</u>isual interactive <u>a</u>nalysis tool for filtering and summarizing large <u>d</u>ata <u>s</u>ets coded with hierarchical terminologies) or other tools.

**Methods:** We recruited clinical researchers and separated them into “experienced” and “inexperienced” groups. Participants were randomly assigned to a VIADS or control group within the groups. Each participant conducted a remote 2-hour study session for hypothesis generation with the same study facilitator on the same datasets by following a think-aloud protocol. Screen activities and audio were recorded, transcribed, coded, and analyzed. Hypotheses were evaluated by seven experts on their validity, significance, and feasibility. We conducted multilevel random effect modeling for statistical tests.

**Results:** Eighteen participants generated 227 hypotheses, of which 147 (65%) were valid. The VIADS and control groups generated a similar number of hypotheses. The VIADS group took a significantly shorter time to generate one hypothesis (e.g., among inexperienced clinical researchers, 258 seconds versus 379 seconds, *p* = 0.046, power = 0.437, ICC = 0.15). The VIADS group received significantly lower ratings than the control group on feasibility and the combination rating of validity, significance, and feasibility.

**Conclusion:** The role of VIADS in hypothesis generation seems inconclusive. The VIADS group took a significantly shorter time to generate each hypothesis. However, the combined validity, significance, and feasibility ratings of their hypotheses were significantly lower. Further characterization of hypotheses, including specifics on how they might be improved, could guide future tool development.

## Highlights of the paper

- Distinguished the scientific hypothesis generation process from other parts of scientific or medical thinking and reasoning.
- Conducted a human participant study to generate data-driven hypotheses among clinical researchers.
- Established baseline data for inexperienced clinical researchers: the number, the quality, the validity rate, and the time needed to generate data-driven hypotheses within 2 hours.
- VIADS seems to facilitate users to generate hypotheses more efficiently.
- Cognitive overload can be a critical factor in negatively influencing the quality of the hypothesis generated.

## Introduction

A scientific hypothesis is an educated guess regarding the relationships among several variables [1,2]. A hypothesis is a fundamental component of a research question [3], which typically can be answered by testing one or several hypotheses [4]. A hypothesis is critical for any research project; it determines its direction and impact. Many studies focusing on scientific research have made significant progress in scientific [5,6] and medical reasoning [7–11], problem-solving, analogy, working memory, and learning and thinking in educational contexts [12]. However, most of these studies begin with a question and focus on scientific reasoning [14], medical diagnosis, or differential diagnosis [10,15,16]. Henry and colleagues named them as open or closed discoveries in the literature-mining context [18]. The reasoning mechanisms and processes used in solving an existing puzzle are critical; however, the current literature provides limited information about the scientific hypothesis generation process [4–6], which is to identify the focused area to start with, not the hypotheses generated to solve existing problems.

There have been attempts to generate hypotheses automatically using text mining, literature mining, knowledge discovery, natural language processing techniques, Semantic Web technology, or machine learning methods to reveal new relationships among diseases, genes, proteins, and conditions [19–23]. Many of these efforts were based on Swanson’s ABC Model [24–26]. Several research teams explored automatic literature systems for generating [28,29] and validating [30] or enriching hypotheses [31]. However, the studies recognized the complexity of the hypothesis generation process and concluded that it does not seem feasible to generate hypotheses completely automatically [19–21,24,32]. In addition, hypothesis generation is not just identifying new relationships although a new connection is a critical component of hypothesis generation. Other literature-related efforts include adding temporal dimensions to machine learning models to predict connections between terms [33,34] or evaluating hypotheses using knowledge bases and Semantic Web technology [32,35]. To understand how humans use such systems to generate hypotheses in practice may provide unique insights into our understanding of scientific hypothesis generation, which can help system developers to better automate systems to facilitate the process.

Many researchers believe that their secondary data analytical tools (such as a <u>v</u>isual interactive <u>a</u>nalytic tool for filtering and summarizing large health <u>d</u>ata <u>s</u>ets coded with hierarchical terminologies —VIADS [36–39]) can facilitate hypothesis generation [40,41]. Whether these tools work as expected, and how, has not been systematically investigated. Data-driven hypothesis generation is critical and the first step in clinical and translational research projects [42]. Therefore, we conducted a study to investigate if and how VIADS can facilitate generating data-driven hypotheses among clinical researchers. We recorded their hypothesis generation process and compared the results of those who used and did not use VIADS. Hypothesis quality evaluation is usually conducted as part of a larger work, e.g., the evaluation of a scientific paper or a research grant proposal. Therefore, there are no existing metrics for hypothesis quality evaluation. We developed our quality metrics to evaluate hypotheses [1,3,4,43–49] through iterative internal and external validation [43,44]. This paper is a study of scientific hypothesis generation by clinical researchers with or without VIADS, including the quality evaluation, quantitative measurement of the hypotheses, and an analysis of the responses to follow-up questions. This is a randomized human participant study; however, per the definition of the National Institutes of Health, it is not a clinical trial. Therefore, we did not register it on ClinicalTrials.gov.

## Methods

### Research question and hypothesis

- Can secondary data analytic tools, e.g., VIADS, facilitate the hypothesis generation process?

We hypothesize there will be group differences between clinical researchers who use VIADS in generating hypotheses and those who do not.

### Rationale of the research question

Many researchers believe that new analytical tools offer opportunities to reveal further insights and new patterns in existing data to facilitate hypothesis generation [1,28,41,42,50]. We developed the underlying algorithms (determine what VIADS can do) [38,39] and the publicly accessible online tool—VIADS [37,51,52] to provide new ways of summarizing, comparing, and visualizing datasets. In this study, we explored the utility of VIADS.

### Study design

We conducted a 2 × 2 study. We divided participants into four groups: inexperienced clinical researchers without VIADS (group 1, participants were free to use any other analytical tools), and with VIADS (group 2). Experienced clinical researchers without VIADS (group 3), and with VIADS (group 4). The main differences between experienced and inexperienced clinical researchers were years of experience in conducting clinical research and the number of publications as significant contributors [53].

A pilot study, involving two participants and four study sessions, was conducted before we finalized the study datasets (Supplemental Material 1), training material (Supplemental Material 2), study scripts (Supplemental Material 3), follow-up surveys (Supplemental Material 4 & 5), and study session flow. Afterward, we recruited clinical researchers for the study sessions.

### Recruitment

We recruited study participants through local, national, and international platforms, including American Medical Informatics Association (AMIA) mailing lists for working groups (e.g., clinical research informatics, clinical information system, implementation, clinical decision support, and Women in AMIA), N3C [54] study network Slack channels, South Carolina Clinical and Translational Research Institute newsletter, guest lectures and invited presentations in peer-reviewed conferences (e.g., MIE 2022), and other internal research related newsletters. All collaborators of the investigation team shared the recruitment invitations with their colleagues. Based on the experience level and our block randomization list, the participants were assigned to the VIADS or non-VIADS groups. After scheduling, the study script and IRB-approved consent forms were shared with participants. The datasets were shared on the study date. All participants received compensation based on the time they spent.

### Study flow

Every study participant used the same datasets and followed the same study scripts. The same study facilitator conducted all study sessions. For the VIADS groups, we scheduled a training session (one-hour). All groups had a study session lasting a maximum of 2 hours. During the training session, the study facilitator demonstrated how to use VIADS and then the participants demonstrated the use of VIADS. During the study session, participants analyzed datasets with VIADS or other tools to develop hypotheses by following the think-aloud protocol. During the study sessions, the study facilitator asked questions, provided reminders, and acted as a colleague to the participants. All training and study sessions were conducted remotely via WebEx meetings. Figure 1 shows the study flow.

**Figure 1.**
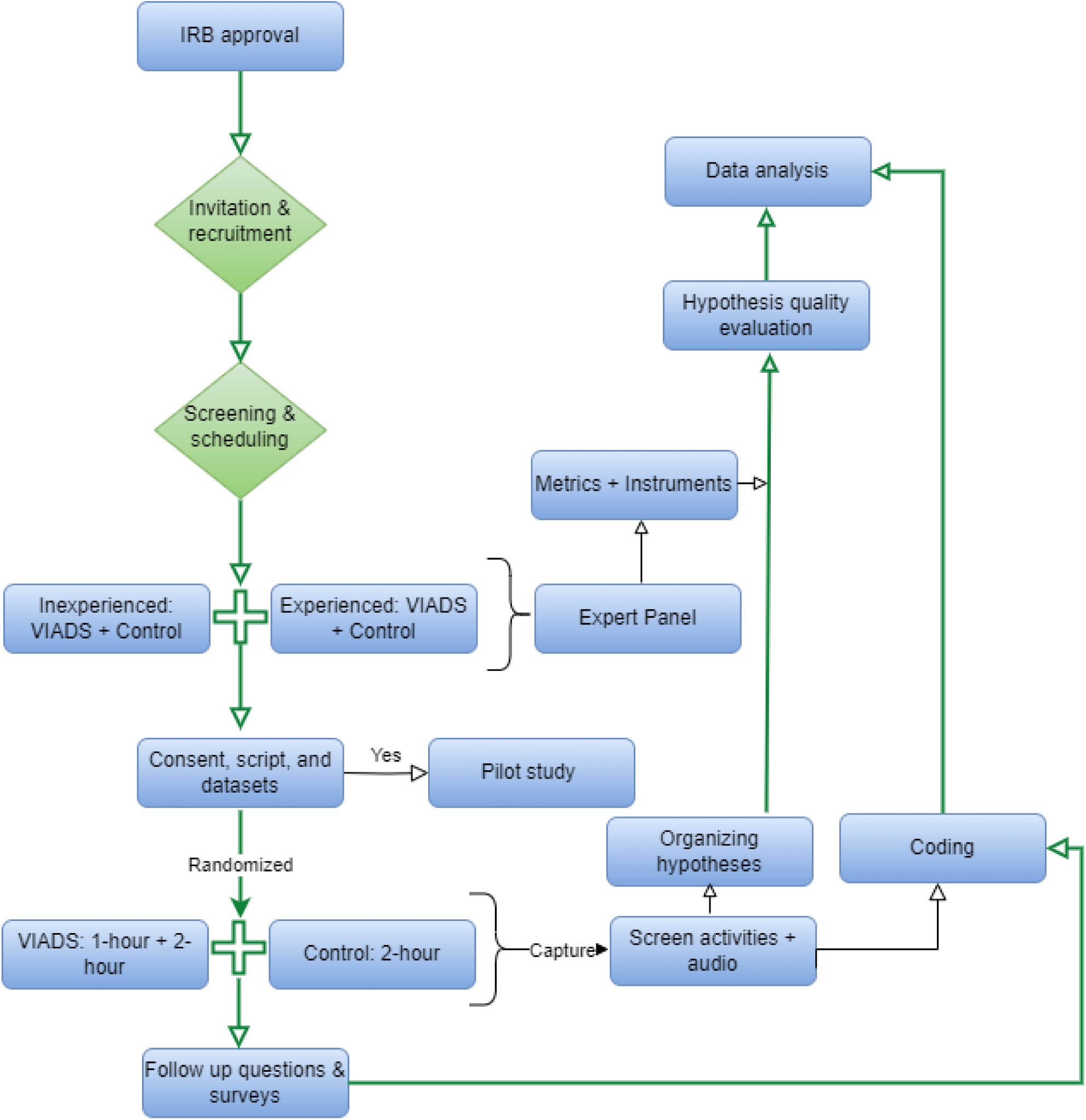
Study flow for the data-driven hypothesis generation.

During the study session, all the screen activities and conversations were recorded via FlashBB and converted to audio files for professional transcription. At the end of each study session, the study facilitator asked follow-up questions about the participants’ experiences creating and capturing new research ideas. The participants in the VIADS groups also completed two follow-up Qualtrics surveys; one was about the participant and questions on how to facilitate the hypothesis generation process better (Supplemental Material 4), and the other evaluated VIADS usability with a modified version of the System Usability Scale (Supplemental Material 5). The participants in the non-VIADS groups received one follow-up Qualtrics survey (Supplemental Material 4).

### Hypothesis evaluation

We developed a complete version and a brief version of hypothesis quality evaluation instrument (Supplemental Material 6) based on hypothesis quality evaluation metrics. We recruited a clinical research expert panel with four external members and three senior project advisors from our investigation team with clinical research backgrounds to validate the instruments. Their detailed eligibility criteria were published [53]. The expert panel evaluated the quality of all hypotheses generated by participants. In Phase 1, the full version of the instrument was used to evaluate randomly selected 30 hypotheses, and the evaluation results enabled us to develop a brief version of the instrument [43] (Supplemental Material 7, including three dimensions: validity, significance, and feasibility); Phase 2 used the brief instrument to evaluate the remaining hypotheses. Each dimension used a 5-point scale, from 1 (the lowest) to 5 (the highest). Therefore, for each hypothesis, the total raw score could range between 3 and 15. In our descriptive results and statistical tests, we used the averages of these scores (i.e., total raw score/3). Like the scores for each dimension, the results ranged from 1 to 5.

We generated a random list for all hypotheses. Then, based on the random list, we put ten randomly selected hypotheses into one Qualtrics survey for quality evaluation. We initiated the quality evaluation process after the completion of all the study sessions, allowing all hypotheses to be included in the generation of the random list.

### Data analysis plan on hypothesis quality evaluation

Our data analysis focuses on the quality and quantity of hypotheses generated by participants. We conducted multilevel random intercept effect modeling in MPlus 7 to compare the VIADS group and the control group on the following items: the quality of the hypotheses, the number of hypotheses, and the time needed to generate each hypothesis by each participant. We also examined the correlations between the hypothesis quality ratings and the participant’s self-perceived creativity.

We first analyzed all hypotheses to explore the aggregated results. A second analysis was conducted by using only valid hypotheses after removing any hypothesis that was scored at “1” (the lowest rating) for validity by three or more experts. However, we include both sets of results in this paper. The usability results of VIADS were published separately [36].

All hypotheses were coded by two research assistants who worked separately and independently. They coded the time needed for each hypothesis and cognitive events during hypothesis generation. The coding principles (Supplemental Material 8) were developed as the two research assistants worked. Whenever there was a discrepancy, a third member of the investigation team joined the discussion to reach a consensus by refining the coding principles.

### Ethical statement

Our study was approved by the Institutional Review Boards (IRB) of Clemson University, South Carolina (IRB2020-056) and Ohio University (18-X-192).

## Results

### Participant demographics

We screened 39 researchers, among whom 20 participated, of which two were in the pilot study. Participants were from different locations and institutions in the United States. Among the 18 study participants, 15 were inexperienced clinical researchers and three were experienced. The experienced clinical researchers were underrepresented, and their results were mainly for informational purposes. Table 1 presents the background information of the participants.

**Table 1.** Eighteen participants’— clinical researchers’ profile.

| Gender | Inexperienced VIADS group | Inexperienced Control group | Gender | Experienced VIADS group | Experienced Control group |
| --- | --- | --- | --- | --- | --- |
| Female | 5 (62.5%) | 4 (57.1%) | - | - | - |
| Male | 3 (37.5%) | 3 (42.9%) | Male | 2 | 1 |
| **Hypothesis generation experience (years)** |  |  |  |  |  |
| < 2 | 6 (75%) | 3 (42.9%) | ≥ 5 and < 10 | 1 | 1 |
| 2–5 | 2 (25%) | 3 (42.9%) | ≥ 10 | 1 | - |
| > 5 | - | 1 (14.3%) | - | - | - |
| Collaborating role | 5 (62.5%) | 5 (71.4%) | Collaborating role | 1 | - |
| Leading role | 1 (12.5%) | 1 (14.3%) | Leading role | 1 | 1 |
| Other | 2 (25%) | 1 (14.3%) | - | - | - |
| **Design experience (years)** |  |  |  |  |  |
| < 2 | 6 (75%) | 3 (42.9%) | ≥ 5 and < 10 | 2 | 1 |
| 2–5 | 2 (25%) | 4 (57.1%) | - | - | - |
| Collaborating role | 6 (75%) | 5 (71.4%) | Collaborating role | 1 | - |
| Leading role | - | 1 (14.3%) | Leading role | 1 | 1 |
| Other | 2 (25%) | 1 (14.3%) | - | - | - |

| Data analysis experience (years) |  |  |  |  |  |
| --- | --- | --- | --- | --- | --- |
| < 2 | 7 (87.5%) | 3 (42.9%) | ≥ 5 and < 10 | 1 | 1 |
| 2–5 | 1 (12.5%) | 4 (57.1%) | ≥ 10 | 1 | - |
| Number of main publications |  |  |  |  |  |
| < 5 | 7 (87.5%) | 7 (100%) | 5–10 | - | 1 |
| ≥ 5 and < 10 | 1 (12.5%) | - | ≥ 10 | 1 | 1 |
| Specialties/Domain |  |  |  |  |  |
| Pediatrics | - | 1 (14.3%) | - | - | - |
| Primary care provider | 1 (12.5%) | 2 (28.6%) | - | - | - |
| Health science | 2 (25%) | 2 (28.6%) | - | - | - |
| Clinical research coordinator | 2 (25%) | 1 (14.3%) | - | - | - |
| Pharmacy | 2 (25%) | - | - | - | - |
| Nursing | - | 1 (14.3%) | - | - | - |
| Internal medicine | 1 (12.5%) | - | - | - | - |
| Routine data analytic tools |  |  |  |  |  |
| MS Excel | 6 | 4 | MS Excel | 1 | 1 |
| R | 3 | 2 | R | - | 1 |
| SPSS | - | 1 | SPSS | 2 | - |
| SAS | 2 | 2 | - | - | - |
| Stata | - | 1 | - | - | - |

Note: -, no value.

### Expert panel composition and intraclass correlation coefficient (ICC)

Seven experts validated the metrics and instruments [43,44] and evaluated the hypotheses using the instruments. Each expert was from a different institution in the United States. Five had medical backgrounds, three of them were in clinical practice; and two had research methodology expertise. They all had 10 years or longer clinical research experience. For the hypothesis quality evaluation, the ICC of the seven experts was moderate, at 0.49.

### Hypothesis quality and quantity evaluation results

The 18 participants generated 227 hypotheses during the study sessions. There were 80 invalid hypotheses and 147 (65%) valid hypotheses. They were all used separately for further analysis and comparison. Of these 147, 121 were generated by inexperienced clinical researchers (n = 15) in the VIADS (n = 8) and control (n = 7) groups.

Table 2 shows the descriptive results of the hypothesis quality evaluation results between the VIADS and the control groups. We used four analytic strategies: valid hypotheses by inexperienced clinical researchers (n = 121), valid hypotheses by inexperienced and experienced clinical researchers (n = 147), all hypotheses by inexperienced clinical researchers (n = 192), and all hypotheses by inexperienced and experienced clinical researchers (n = 227).

**Table 2.** Expert panel quality rating results for hypotheses generated by VIADS and control groups.

| Analytic strategies | Seven expert rating hypothesis group mean |  |  |  |  |  |  |  |
| --- | --- | --- | --- | --- | --- | --- | --- | --- |
|  | Overall/3 |  | Validity |  | Significance |  | Feasibility |  |
|  | VIADS Mean (SD) | Control Mean (SD) | VIADS Mean (SD) | Control Mean (SD) | VIADS Mean (SD) | Control Mean (SD) | VIADS Mean (SD) | Control Mean (SD) |
| Inexperienced + valid. (121) | 2.97 (0.45) | 3.17 (0.45) | 2.79 (0.59) | 2.85 (0.56) | 3.14 (0.52) | 3.15 (0.57) | 2.98 (0.77) | 3.53 (0.77) |
| Inexperienced + experienced + valid. (147) | 2.93 (0.45) | 3.19 (0.43) | 2.75 (0.49) | 2.86 (0.54) | 3.17 (0.49) | 3.19 (0.54) | 2.88 (0.76) | 3.52 (0.74) |
| Inexperienced + all. (192) | 2.81 (0.51) | 3.01 (0.51) | 2.45 (0.59) | 2.56 (0.59) | 3.09 (0.60) | 3.14 (0.60) | 2.86 (0.83) | 3.33 (0.83) |
| Inexperienced + experienced + all. (227) | 2.79 (0.53) | 3.05 (0.53) | 2.43 (0.59) | 2.60 (0.62) | 3.10 (0.57) | 3.18 (0.60) | 2.80 (0.81) | 3.33 (0.86) |

**Note:** valid. → valid hypotheses; all. → all hypotheses; SD, standard deviation. Each hypothesis was rated on a 5-point scale, from 1 (the lowest) to 5 (the highest) in each dimension.

Table 3 shows the hypotheses’ quality evaluation results for random intercept effect modeling. The four analytic strategies generated similar results. The VIADS group received slightly lower validity, significance, and feasibility scores, but the differences in validity and significance were statistically insignificant regardless of the analytic strategy. However, the feasibility scores of the VIADS group were statistically significantly lower (*p* < 0.01), regardless of the analytic strategy. Using the random intercept effect model, the combined validity, significance, and feasibility ratings of the VIADS group were statistically significantly lower than those of the control group for three of four analytic strategies (*p* < 0.05). This was most likely due to the differences in feasibility ratings between the VIADS and control groups. Across all four analytic strategies, there was a statistically significant random intercept effect on validity between participants. When all hypotheses and all participants were considered, there was also a statistically significant random intercept effect on significance between participants.

**Table 3.** Multilevel random intercept modeling results on hypotheses quality ratings for different strategies.

| Analytic strategy | Parti. | Power | Overall/3 |  |  | Validity |  |  | Significance |  |  | Feasibility |  |  |
| --- | --- | --- | --- | --- | --- | --- | --- | --- | --- | --- | --- | --- | --- | --- |
| | | | $\beta$ | $p$ | ICC | $\beta$ | $p$ | ICC | $\beta$ | $p$ | ICC | $\beta$ | $p$ | ICC |
| Inexperienced + valid. (121) | 15 | 0.37 | -0.21 | 0.02 | 0.06 | -0.09 | 0.51 | 0.2 | -0.03 | 0.82 | 0.08 | -0.53 | <0.01 | 0.01 |
| Inexperienced + experienced + valid. (147) | 18 | 0.55 | -0.26 | <0.01 | 0.06 | -0.14 | 0.36 | 0.21 | -0.02 | 0.89 | 0.09 | -0.63 | <0.001 | 0.03 |
| Inexperienced + all. (192) | 15 | 0.58 | -0.2 | 0.06 | 0.06 | -0.14 | 0.44 | 0.26 | -0.05 | 0.66 | 0.07 | -0.44 | <0.01 | 0.02 |
| Inexperienced + experienced + all. (227) | 18 | 0.79 | -0.26 | 0.01 | 0.08 | -0.22 | 0.20 | 0.26 | -0.09 | 0.43 | 0.09 | -0.53 | <0.001 | 0.04 |

**Note:** valid. → valid hypotheses; all. → all hypotheses; Parti. → participants; ICC: intraclass correlation coefficient. $\beta$ , group mean difference between VIADS and control groups in their quality ratings of hypotheses; ICC (for random effect modeling) = between-participants variance/(between-participants variance + within-participants variance).

In addition to quality ratings, we also compared the number of hypotheses and the time needed for each participant to generate a hypothesis between groups. The inexperienced clinical researchers in the VIADS group and control group generated a similar number of valid hypotheses. Those in the VIADS group generated between 1 and 19 valid hypotheses in 2 hours (mean 8.43) and those in the control group generated 2 to 13 (mean 7.63). The inexperienced clinical researchers in the VIADS group took 102–610 seconds to generate a valid hypothesis (mean, 278.6 s), and those in the control group took 250–566 seconds (mean, 358.2 s).

Table 4 shows the random intercept modeling results of our comparison of the time needed to generate hypotheses using the four different strategies. On average, the VIADS group requires significantly less time (*p* < 0.05) to generate a hypothesis regardless of the analytic strategy used. The results were consistent with those obtained from the analysis of all hypotheses by independent t-test [55]. There were no statistically significant random intercept effects between participants, regardless of the analytic strategy used (Table 4).

**Table 4.** Multilevel random intercept modeling results on time used to generate hypotheses for different strategies.

| Analytic strategy | Parti. | Power | $\beta$ | $p$ | ICC |
| --- | --- | --- | --- | --- | --- |
| Inexperienced + valid. (121) | 15 | 0.44 | -119.45 | 0.046 | 0.15 |
| Inexperienced + experienced + valid. (147) | 18 | 0.43 | -124.81 | 0.02 | 0.12 |
| Inexperienced + all. (192) | 15 | 0.32 | -97.93 | 0.02 | 0.11 |
| Inexperienced + experienced + all. (227) | 18 | 0.31 | -95.51 | 0.02 | 0.12 |

**Note:** valid. → valid hypotheses; all. → all hypotheses; Parti. → participants; ICC: intraclass correlation coefficient; $\beta$ , group mean difference between VIADS and control groups in the time used to generate each hypothesis on average; ICC (for random effect modeling) = between-participants variance/(between-participants variance + within-participants variance)

### Experienced clinical researchers

There were three experienced clinical researchers among the participants, two in the VIADS group and one in the control group. The experienced clinical researchers in the VIADS group generated 12 (average time 215 s/hypothesis) and 3 (average time 407 s/hypothesis) valid hypotheses in 2 hours. The experienced clinical researcher in the control group generated 12 valid hypotheses (average time, 413 s/hypothesis). The experienced clinical researchers in the VIADS group received average quality scores of 8.51 (SD, 1.18) and 7.04 (SD, 0.3) while the experienced clinical researcher in the control group received a quality score of 9.84 (SD, 0.81) out of 15 per hypothesis. These results were used for informational purpose.

### Follow-up questions

The follow-up questions comprise three parts: verbal questions asked by the study facilitator and a follow-up survey for all participants, and a SUS usability survey for the VIADS group participants [36]. The results from the first two parts are summarized below.

The verbal questions and the summary answers are presented in Table 5. Reading and interactions with others were the most used activities to generate new research ideas. Attending conferences, seminars, and educational events, and conducting clinical practice were important in generating hypotheses. There were no specific tools used to initially capture hypotheses or research ideas. Most participants used text documents in Microsoft Word, text messages, emails, or sticky notes to summarize their initial ideas.

**Table 5.**
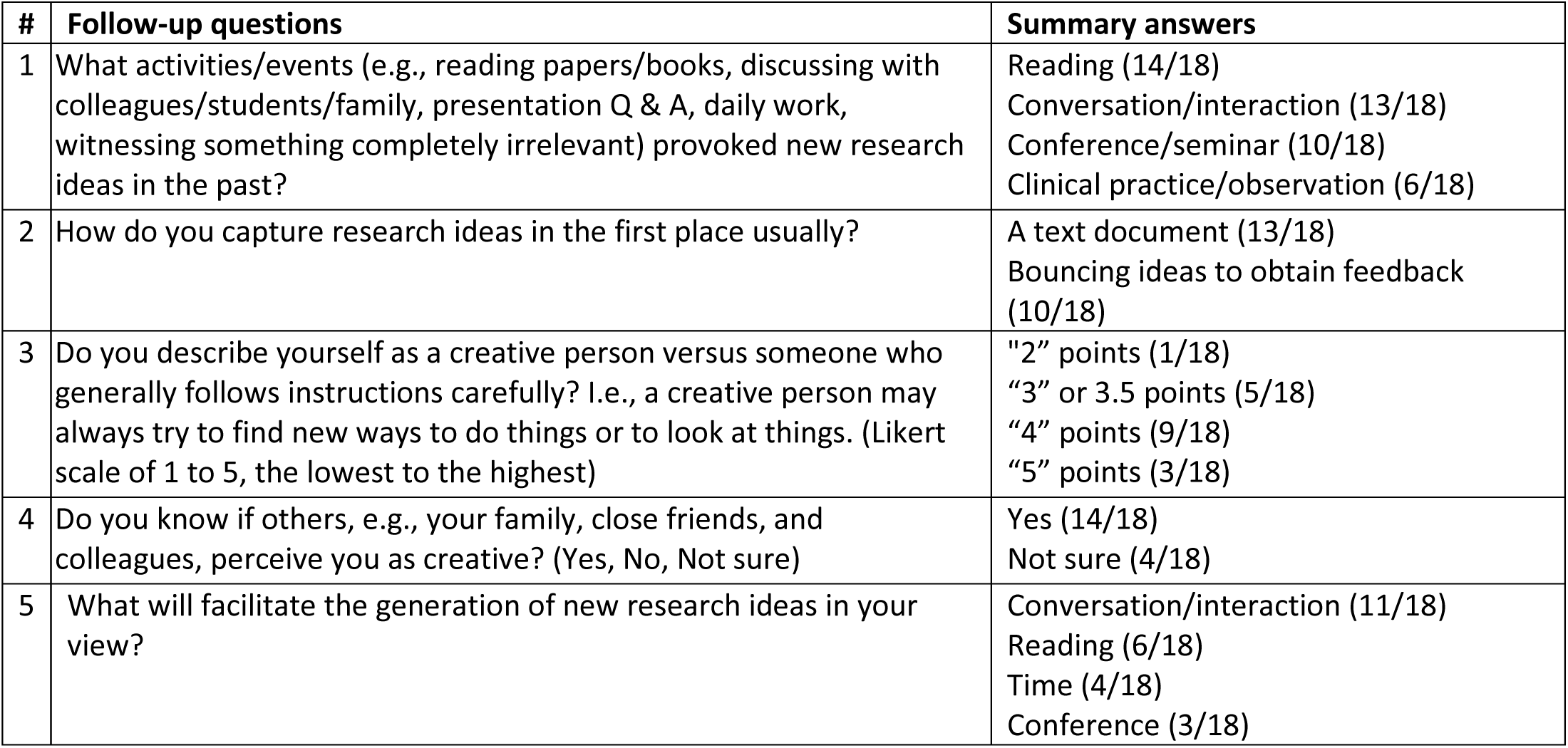
Follow-up questions (verbal) and answers after each study session (all study participants)

Figure 2 is a scientific hypothesis generation framework we developed based on literature [1,3,4,45,56,57], follow-up questions and answers after study sessions, and self-reflection on our research project trajectories. The external environment, cognitive capacity, and interactions between the individual and the external world, especially the tools used, are categories of critical factors that significantly contribute to hypothesis generation. Cognitive capacity takes a long time to change, and the external environment can be unpredictable. The tools that can interact with existing datasets are one of the modifiable factors in the hypothesis generation framework and this is what we aimed to test in this study.

**Figure 2.**
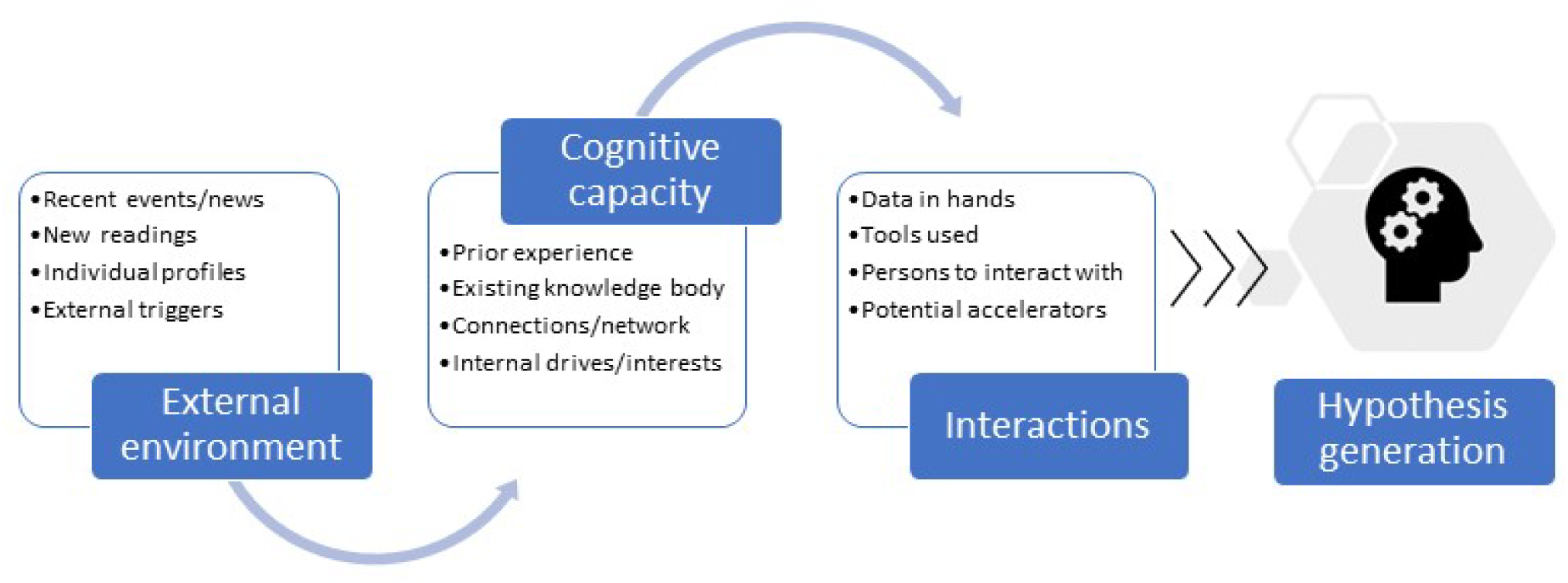
Scientific hypothesis generation framework: Contributing factors.

One follow-up question was about the self-perceived creativity of the study participants. The average hypothesis quality rating score per participant did not correlate with the self-perceived creativity (*p* = 0.616, two-tailed Pearson correlation test) or the number of valid hypotheses generated (*p* = 0.683, two-tailed Pearson correlation test) by inexperienced clinical researchers. There was no correlation between the highest and lowest 10 ratings and the individual’s self-perceived creativity, in inexperienced clinical researchers regardless of using VIADS or not.

In our follow-up survey, the questions were mainly about participants’ current roles and affiliations, their experience in clinical research, their preference for analytic tools, and their ratings of the importance of different factors (e.g., prospective study, longitudinal study, and the results will be published separately) considered routinely in clinical research study design. Most of the results have been included in Table 1.

In our follow-up survey, one question was, “If you were provided with more detailed information about research design (e.g., focused population) during your hypothesis generation process, do you think the information would help formulate your hypothesis overall?” All 20 participants, (including two in the pilot study), selected Yes. This demonstrates the recognition and need for assistance during hypothesis generation. In the follow-up surveys, VIADS users provided overwhelmingly positive feedback on VIADS, and they all agreed (100%) that VIADS offered new perspectives on the datasets compared with the tools they currently use for the same type of datasets [36].

## Discussion

### Interpretation of the results

We aim to discover the role of secondary data analytic tools, e.g., VIADS, in generating scientific hypotheses in clinical research and to evaluate the utility and usability of VIADS. The usability of VIADS has been published separately [36]. Regarding the role and utility of VIADS, we measured the number of hypotheses generated, the average time needed to generate each hypothesis, the quality evaluation of the hypotheses, and the user feedback on VIADS. Participants in the VIADS and control groups generated similar numbers of valid and total hypotheses among inexperienced clinical researchers. The VIADS group a) needed a significantly shorter time to generate each hypothesis on average; b) received significantly lower ratings on quality in feasibility than the control group; c) received significantly lower quality ratings (three of four analytic strategies) in the combination ratings of validity, significance, and feasibility, which most likely due to the feasibility rating differences; d) provided very positive feedback on VIADS [36] with 75% agreed that VIADS facilitates understanding, presentation, and interpretation of the underlying datasets; e) agreed (100%) that VIADS provided new perspectives on the datasets compares to other tools.

However, the current results were inconclusive in answering the research question. The direct measurements of significant differences between the VIADS and control group were mixed (Tables 3 and 4). The VIADS group took significantly less time than the control group to generate each hypothesis, regardless of the analytic strategy. Considering the sample size and power (0.31 to 0.44) of the study (Table 4), and the absence of significant random intercept effects on feasibility between participants, regardless of analytic strategy, this result is highly significant. The shorter hypothesis generation time in the VIADS group indicates that VIADS may facilitate participants’ thinking or understanding of the underlying datasets. While timing is not as critical in the context of clinical research as it is in clinical care, this result is still very encouraging.

On the other hand, the quality ratings of the hypotheses generated mixed and somewhat unfavorable results. The VIADS group received insignificantly lower ratings for validity and significance, and significant lower ratings for feasibility than the control group, regardless of analytic strategy. In addition, the combined validity, significance, and feasibility ratings of the VIADS group were significantly lower than those of the control group for three analytic strategies (the power ranged from 0.37 to 0.79, Table 3). There were significant random intercept effects on validity between participants, regardless of analytic strategy. When we considered all hypotheses among all participants, there were also significant random intercept effects on significance between participants. These results indicate that the significantly lower ratings for feasibility in the VIADS groups may not be caused by random effects. There are various possible reasons for the lower feasibility ratings of the VIADS group, e.g., VIADS facilitates the generation of less feasible hypotheses. While unfeasible ideas are not identical to creative ideas, they may be a deviation on the path to creative ideas.

Although the VIADS group received lower quality ratings than the control group, it would be an overstatement to claim that VIADS reduces the quality of generated hypotheses. We posit that the 1-hour training session received by the VIADS group likely played a critical role in the quality rating differences. Among the inexperienced clinical researchers, six underwent 3-hour sessions, i.e., 1-hour training followed by a 2-hour study session, with a 5-minute break in between. Two participants had the training and the study session on separate days. The cognitive load of the training session was not considered during the study design, therefore the training sessions were not required to be conducted on different days to the study sessions. However, in retrospect, we should have mandated that the training and study sessions take place on separate days. In addition, the VIADS group had a much higher percentage of participants with less than 2 years of research experience (75%) than the control group (43%, Table 1). Although we adhered strictly to the randomization list when assigning participants to the two groups, the relatively small sample sizes are likely to have amplified the effects of the research experience imbalance between the two groups.

Literature [58] suggests that learning a complex tool and performing tasks simultaneously presents extra cognitive load on the participants. This is likely the case in this study. VIADS group participants needed to learn how to use the tool and then analyze the data sets with it to come up with the hypotheses. The cognitive overload may not have been conscious. Therefore, the participants perceived VIADS as helpful in understanding datasets. However, the quality evaluation results did not support the participants’ perceptions of VIADS although the timing differences did support participants’ feedback.

The role of VIADS in the hypothesis generation process may not be linear. The 2-hour use of VIADS did not generate statistically higher average quality ratings on hypotheses; however, all participants (100%) agreed that VIADS provided new perspectives on the datasets. The true role of VIADS in hypothesis generation might be more complicated than we initially thought. Either two hours were inadequate to generate higher average quality ratings, or our evaluation was not adequately granular to capture the differences. A more natural use environment might be necessary instead of a simulated environment to demonstrate detectable differences.

Researchers have long been keen to understand where good research ideas come from [59,60]. Participants’ answers to our follow-up questions have provided anecdotal information about possible activities contributing to the hypothesis generation process. From these insights, and a literature review [1,3,4,45,56,57], we formulated a scientific hypothesis generation framework (Figure 2). All the following activities were identified as associated with hypothesis generation: reading, interactions with others, observations during clinical practice, teaching, learning, and listening to presentations. Individuals connected ideas, facts, and phenomena, and formulated them into research questions and hypotheses to test. Although these activities did not answer the question directly, they were identified by the participants as associated with hypothesis generation.

Before the expert panel members evaluated the hypotheses, it was necessary to establish some form of threshold based on the following considerations. First, most of the participants were inexperienced clinical researchers. Second, the participants were unfamiliar with the data-driven hypothesis generation process. And third, the number of hypotheses each participant generated within a 2-hour window. It was unlikely that each hypothesis generated would be valid or valuable and should be given equal attention by the expert panel. Therefore, if three or more experts rated the validity of a hypothesis as 1 (the lowest), then the hypothesis was considered invalid. However, these hypotheses were included in two of the analytic strategies. Meanwhile, we do recognize that identifying an impactful research question is necessary but does not guarantee a successful research project, but only an excellent start.

Three participants with the highest average quality ratings (i.e., 10.55, 10.25, and 9.84 out of 15) were all in the non-VIADS group, of which the top two were inexperienced and the third was an experienced clinical researcher. They all practice medicine. Based on the conversations between them and the study facilitator, they all put much thought into research and connect observations in medical practice and clinical research; their clinical practice experience, education, observation, thinking, and making connections contributed to their higher quality ratings on their hypotheses. This observation verifies the belief that good research ideas require at least three pillars: 1) a deep understanding of the domain knowledge and the underneath mechanisms, 2) capable of connecting knowledge and practical observations (problems or phenomena), 3) capable to put the observations into the appropriate research contexts [3,4,59,61–63].

The three participants with the highest average quality scores were in the non-VIADS group, and the participant with the lowest average quality score was in the VIADS group. Regardless of our randomization, individual variations may play an amplified role when the sample size is relatively small. Although the random effect models allowed more precise statistical analyses, larger sample sizes would have higher power and more reliable results.

### Significance of the work

Using the same datasets, we conducted the first human participant investigation to compare data-driven hypothesis generation using VIADS and other analytic tools. The significance of the study can be demonstrated in the following aspects. ***First***, this study demonstrated the feasibility of remotely conducting a data-driven hypothesis generation study via the think-aloud protocol. ***Second***, we established hypothesis quality evaluation metrics and instruments, which may be useful for clinical researchers during peer review or prioritize their research ideas before investing too many resources. ***Third***, this study measured the baseline data for hypotheses, time needed, and quality of hypotheses generated by inexperienced clinical researchers. ***Fourth***, hypothesis generation is complicated, and our results showed the VIADS group generated hypotheses significantly faster than the control group, and the overall quality of the hypothesis was favorable in the control group. ***Fifth***, among inexperienced clinical researchers, we identified that more assistance is needed in the hypothesis generation process. This work has laid a foundation for more structured and organized clinical research projects, starting from a more explicit hypothesis generation process. However, we believe that this is only the tip of the hypothesis generation iceberg.

### Strengths and limitations of the work

The study participants were from all over the United States, not a single health system or institution. Although the sample of participants may be more representative, individual variations may have played a more significant role than estimated in the hypothesis quality measurements.

We implemented several strategies to create comparable groups. For example, we used the same datasets, study scripts, and platform (WebEx) for all study sessions conducted by the same study facilitator. Two research assistants examined the time measurements and events of the hypothesis generation process independently and compared their results later, which made the coding more consistent and robust.

The study had a robust design and a thorough analysis. Random intercept effect modeling was a more precise statistical test. We implemented randomization at multiple levels to reduce potential bias. The study participants were separated into experienced and inexperienced groups during screening based on predetermined criteria [53] and then randomly assigned to the VIADS or non-VIADS groups. For the hypothesis quality evaluation, every ten randomly selected hypotheses were organized into one Qualtrics survey. This provided a fair opportunity for each hypothesis and reduced bias related to the order of hypotheses. The hypothesis quality measurement metrics and instruments were validated iteratively, internally, and externally [43]. We implemented multiple analytical strategies to provide a comprehensive picture of the data and enhance the robustness of the study. We examined (a) valid hypotheses only; (b) inexperienced clinical researchers only; (c) all hypotheses; and (d) all clinical researchers.

One major limitation was the sample size, which was based on our ability to recruit over the study period and was insufficient to detect an effect of the size we originally anticipated. The power calculation (i.e., the required sample size was 32 for four groups based on the confidence level of 95%, *α* = 0.05, and a power level of 0.8, *β* = 0.20) used during the study design was optimistic, mainly due to our belief that VIADS can provide new ways of data visualization and data analysis than other tools. The VIADS groups participants verified our confidence in VIADS via the follow-up surveys. However, the quantitative measures showed mixed results that only partially support our opinion and the real power ranged between 0.31 to 0.79. One possible explanation is that hypothesis generation is highly complicated. A tool like VIADS may be helpful, but the effects might not be easily measured or detected after 2–3 hours of use. Better control of participant characteristics would have reduced the possibility of bias due to participant variations between the two groups. In practice, there is no universally agreed-upon measure of research expertise that could be adopted in our study.

Another limitation is that we used a simulated environment. There is a time constraint during study sessions. Time pressure can reduce hypothesis generation [64]. However, this influence would be similar in both groups. The hypothesis generation process in real life is usually lengthy and non-linear, with discussions, revisions, feedback, and refinement. We do not know whether a simulated environment can reflect the true natural process of hypothesis generation. The study facilitator who conducted the study sessions could have been a stressor for some participants.

Our current measurements may be inadequate to detect such complexity. All the VIADS group participants agreed that VIADS is helpful in hypothesis generation; however, the quantitative and qualitative measures presented mixed results, which may indicate more granular measurements are needed. We did notice that hypotheses generated by VIADS groups seem more complex than the control groups, however, we did not implement a measure for complexity of hypothesis to provide concrete evidence.

The ICC for the quality ratings of the hypotheses by seven experts was moderate, at 0.49, which is understandable, considering a hypothesis provides limited information in two to three sentences. In contrast, reviewers often disagree on a full paper or a grant proposal, which include much more details. Despite these points, we still would like to emphasize the complex nature of this study. We should keep an open mind while interpreting the results.

### Challenges and future directions

The cognitive events and processes during hypothesis generation (i.e., the think-aloud recordings of study sessions) are beyond the scope of this paper but are currently being analyzed, to be published separately. The results of this analysis may shed more light on our outcomes. In this study, we faced several challenges beyond our control that may have affected the study results. The current process can only capture the conscious and verbalized processes and may, for example, have failed to capture unconscious cognitive processes. Therefore, further analysis of the recorded think-aloud sessions might help us to better understand the hypothesis generation process and the differences between groups. A large-scale replication of the present study with more participants is needed. The factors that influence the validity, significance, and feasibility of hypotheses should also be explored further.

The VIADS group participants provided very positive feedback on VIADS and its role in hypothesis generation; however, the limitations of VIADS are apparent. By nature, VIADS is a data analysis and visualization tool. It can only accept specific dataset format and only supports certain types of hypotheses. Therefore, more powerful and comprehensive tools designed to assist hypothesis generation particularly are needed [60]. Furthermore, a longer duration of use and use of the tool in a more natural environment might be necessary to demonstrate the tools’ effectiveness.

Recruitment is always challenging in human participant studies [47–49]. It is particularly challenging to recruit experienced clinical researchers, even though we made similar efforts and used similar platforms to recruit inexperienced and experienced clinical researchers. The different recruitment outcomes may be because hypothesis generation is not a high priority for experienced clinical researchers. It could also be because they were overwhelmed by existing responsibilities and did not have time for the additional tasks needed by our study.

## Conclusion

The role of VIADS in hypothesis generation for clinical research seems uncertain. The VIADS group took significantly less time to generate hypotheses than the control group. However, the overall quality (as measured by the combination of validity, significance, and feasibility scores) of the hypotheses generated in the VIADS group was significantly lower than those of the control group; although, there were statistically significant random intercept effects on validity between participants, regardless of the analytic strategy used. The lower combined quality ratings were likely due to the significantly lower feasibility ratings of the VIADS group. Further characterization of hypotheses, including specific ways in which they might be improved, could guide future tool development. Larger-scale studies may help to reveal more conclusive hypothesis generation mechanisms.

## Supporting information

the study datasets (Supplemental Material 1)

training material (Supplemental Material 2),

study scripts (Supplemental Material 3),

follow-up surveys (Supplemental Material 4 & 5),

follow-up surveys (Supplemental Material 4 & 5),

hypothesis quality evaluation instrument (Supplemental Material 6)

develop a brief version of the instrument [43] (Supplemental Material 7,

The coding principles (Supplemental Material 8)

## Data Availability

All data produced in the present study are available upon reasonable request to the authors on a case by case basis.

## Acknowledgment

N/A

## Supplemental Materials

Supplemental Material 1: The data sets used during the hypothesis generation study

Supplemental Material 2: Training materials used for participants in VIADS groups

Supplemental Material 3: Study scripts for participants in VIADS and non-VIADS groups

Supplemental Material 4: Follow-up survey after the hypothesis generation study

Supplemental Material 5: Modified version of System Usability Scale

Supplemental Material 6: Hypothesis quality evaluation instrument for clinical research—full version

Supplemental Material 7: Hypothesis quality evaluation instrument for clinical research—brief version

Supplemental Material 8: Coding principles for timing the hypothesis generation process

