## Supplementary material for "Data-driven hypothesis generation among inexperienced clinical researchers: A comparison of secondary data analyses with visualization (VIADS) and other tools": the study datasets (Supplemental Material 1)

| NAMCS2005_Top137NC | |
|---|---|
| ICD9 codes | Frequency |
| 4019 | 1733 |
| V6759 | 957 |
| V202 | 866 |
| 25000 | 688 |
| 2724 | 635 |
| 4659 | 535 |
| V5889 | 461 |
| V6709 | 453 |
| 4739 | 412 |
| 311 | 404 |
| 4779 | 397 |
| 41400 | 378 |
| V7231 | 366 |
| V4589 | 363 |
| 3829 | 348 |
| 3669 | 331 |
| V221 | 325 |
| 30000 | 319 |
| 53081 | 312 |
| 49390 | 281 |
| 2720 | 278 |
| V700 | 277 |
| 29620 | 265 |
| 7020 | 265 |
| 462 | 260 |
| 6929 | 256 |
| 2449 | 232 |
| 5990 | 219 |
| 31401 | 217 |
| 7061 | 212 |
| 60000 | 208 |
| 34690 | 204 |
| 7242 | 203 |
| 496 | 200 |
| 27800 | 199 |
| 490 | 197 |
| V6549 | 191 |
| 7840 | 190 |
| 2169 | 189 |
| 71590 | 184 |
| 185 | 183 |
| 3004 | 183 |
| 42731 | 166 |
| 29680 | 162 |
| 7862 | 153 |

| | |
|---|---|
| 3659 | 151 |
| 78039 | 150 |
| 78650 | 147 |
| V990 | 147 |
| V508 | 145 |
| 7245 | 144 |
| 3804 | 142 |
| 4720 | 141 |
| 78079 | 141 |
| V7189 | 141 |
| 1739 | 138 |
| 4280 | 132 |
| 5997 | 131 |
| 71596 | 129 |
| 3671 | 128 |
| 7291 | 128 |
| 7804 | 127 |
| 3540 | 125 |
| 7999 | 125 |
| 2859 | 123 |
| 1749 | 120 |
| 7810 | 120 |
| 470 | 119 |
| 71946 | 119 |
| 70219 | 116 |
| 31400 | 115 |
| 78057 | 115 |
| 7231 | 114 |
| 30001 | 113 |
| 7295 | 112 |
| 78609 | 111 |
| 79093 | 110 |
| 3320 | 106 |
| 5920 | 106 |
| 7062 | 102 |
| 30002 | 101 |
| 4619 | 101 |
| 78900 | 97 |
| V7647 | 95 |
| 340 | 94 |
| V431 | 94 |
| 4660 | 93 |
| V720 | 93 |
| 73300 | 92 |
| 4781 | 90 |
| 38181 | 88 |
| V1083 | 88 |

| | |
|---|---|
| 71941 | 87 |
| 25050 | 86 |
| 3003 | 84 |
| 56400 | 84 |
| 71690 | 83 |
| 7820 | 83 |
| 38910 | 82 |
| 78820 | 82 |
| 36500 | 79 |
| 38010 | 79 |
| 7823 | 79 |
| 7140 | 78 |
| V997 | 78 |
| 37230 | 77 |
| 6953 | 77 |
| 340 | 76 |
| 30981 | 76 |
| 8470 | 76 |
| V5883 | 76 |
| V1249 | 75 |
| 7244 | 74 |
| 7962 | 74 |
| 29630 | 73 |
| 5939 | 73 |
| 4439 | 72 |
| 36250 | 71 |
| 78052 | 70 |
| 3674 | 68 |
| 4011 | 68 |
| 4241 | 67 |
| 61610 | 67 |
| 6272 | 67 |
| 7099 | 66 |
| 4139 | 65 |
| 6961 | 65 |
| 37921 | 64 |
| 43491 | 64 |
| 55090 | 64 |
| 7234 | 64 |
| 7821 | 64 |
| 78830 | 64 |
| 486 | 63 |
| 3559 | 61 |
| 3569 | 61 |
| V583 | 61 |

| NAMCS2015_Top125NC | |
|---|---|
| **ICD9 code** | **Frequency** |
| 4019 | 2070 |
| 25000 | 1099 |
| 2724 | 1097 |
| V202 | 924 |
| V5889 | 697 |
| V6759 | 653 |
| V700 | 565 |
| 53081 | 545 |
| 7020 | 538 |
| 30000 | 523 |
| 311 | 513 |
| 70219 | 499 |
| V431 | 499 |
| 41400 | 464 |
| V7231 | 457 |
| 42731 | 442 |
| 2169 | 420 |
| 36616 | 405 |
| 27800 | 404 |
| 7061 | 401 |
| 4779 | 389 |
| 4011 | 384 |
| 2449 | 382 |
| 2720 | 382 |
| V221 | 373 |
| 6929 | 357 |
| 37515 | 350 |
| 49390 | 348 |
| 3669 | 341 |
| V6709 | 340 |
| 4659 | 328 |
| 185 | 325 |
| 3804 | 322 |
| V4589 | 306 |
| 7242 | 306 |
| 31401 | 297 |
| 60001 | 290 |
| 3829 | 286 |
| 7840 | 285 |
| 60784 | 284 |
| 71946 | 282 |
| 4739 | 278 |
| 5990 | 272 |
| 79093 | 257 |
| 2382 | 251 |

| | |
|---|---|
| V6700 | 245 |
| 5920 | 243 |
| 60000 | 236 |
| 38181 | 231 |
| 36570 | 231 |
| 71690 | 226 |
| 70909 | 213 |
| V990 | 212 |
| 7862 | 211 |
| 7295 | 210 |
| 496 | 206 |
| 36511 | 206 |
| 33829 | 204 |
| 7804 | 203 |
| V1083 | 200 |
| 78079 | 200 |
| 7810 | 194 |
| 71596 | 192 |
| 3051 | 191 |
| 470 | 188 |
| 71941 | 187 |
| 2689 | 185 |
| 7245 | 183 |
| 25050 | 182 |
| 37921 | 180 |
| 7062 | 178 |
| 3899 | 176 |
| 34690 | 176 |
| 78841 | 174 |
| 71590 | 172 |
| 32723 | 171 |
| 462 | 167 |
| 4720 | 161 |
| 30002 | 161 |
| 7231 | 157 |
| V508 | 156 |
| 2722 | 156 |
| 3671 | 155 |
| 47819 | 155 |
| 78650 | 154 |
| 3659 | 152 |
| 41401 | 150 |
| 36500 | 149 |
| 2859 | 146 |
| V679 | 144 |
| 78052 | 144 |
| 78900 | 143 |

| | |
|---|---|
| 340 | 137 |
| V2389 | 137 |
| 27801 | 136 |
| V5869 | 136 |
| 1749 | 136 |
| 7291 | 135 |
| 78820 | 134 |
| 78605 | 133 |
| 78609 | 132 |
| 3540 | 132 |
| 7244 | 132 |
| 6953 | 129 |
| 17391 | 129 |
| 7823 | 126 |
| 7820 | 126 |
| 40390 | 125 |
| 29620 | 125 |
| 2572 | 124 |
| 70211 | 123 |
| 6961 | 122 |
| 34590 | 117 |
| 59970 | 116 |
| 29680 | 116 |
| 7099 | 115 |
| 7851 | 115 |
| 6259 | 114 |
| 78830 | 113 |
| 38910 | 113 |
| 4254 | 113 |
| 31400 | 113 |
| 4280 | 111 |
| 5859 | 110 |
| V4561 | 110 |

| ICD9 codes | Abbreviations | Full names |
|---|---|---|
| 1739 | Skin: site unspecified | Skin: site unspecified |
| 17391 | other malignant neoplasm of skin unspecified | other malignant neoplasm of skin: site unspecified |
| 1749 | malignant neoplasm of female breast: unspecified | malignant neoplasm of female breast: unspecified |
| 185 | malignant neoplasm of prostate | malignant neoplasm of prostate |
| 2169 | skin benign neoplasm unspecified | skin benign neoplasm: unspecified site |
| 2382 | neoplasms of uncertain behavior of skin | neoplasms of uncertain behavior of skin |
| 2449 | unspecified hypothyroidism | unspecified hypothyroidism |
| 2500 | diabetes no complication | Diabetes mellitus without mention of complication |
| 25000 | diabetes no complication | diabetes mellitus without mention of complication: type 2 or unspecified type-not stated as uncontrolled |
| 2505 | Diabetes with ophthalmic manifestations | Diabetes with ophthalmic manifestations |
| 25050 | diabetes with ophthalmic manifestations | diabetes with ophthalmic manifestations type 2 or unspecified type not stated as uncontrolled |
| 2572 | other testicular hypofunction | other testicular hypofunction |
| 2689 | unspecified vitamin D deficiency | unspecified vitamin D deficiency |
| 2720 | pure hypercholesterolemia | pure hypercholesterolemia |
| 2722 | mixed hyperlipidemia | mixed hyperlipidemia |
| 2724 | other hyperlipidemia | other and unspecified hyperlipidemia |
| 2780 | Overweight and obesity | Overweight and obesity |
| 27800 | unspecified obesity | unspecified obesity |
| 27801 | morbid obesity | morbid obesity |
| 2859 | unspecified anemia | unspecified anemia |
| 2962 | | Major depressive disorder-single episode |
| 29620 | major depressive disorder: single episode unspecified | major depressive disorder: single episode unspecified |
| 29630 | | Major depressive disorder: recurrent episode unspecified |
| 2968 | | Other and unspecified bipolar disorders |
| 29680 | bipolar disorder: unspecified | bipolar disorder: unspecified |
| 3000 | Anxiety states | Anxiety states |
| 30000 | anxiety state unspecified | anxiety state: unspecified |
| 30001 | | Panic disorder without agoraphobia |
| 30002 | generalized anxiety disorder | generalized anxiety disorder |

| | | |
|---|---|---|
| 3003 | | Obsessive-compulsive disorders |
| 3004 | | Dysthymic disorder |
| 3051 | tobacco use disorder | tobacco use disorder |
| 30981 | | Posttraumatic stress disorder |
| 311 | depression NEC | depressive disorder: not elsewhere classified |
| 3140 | Attention deficit disorder | Attention deficit disorder |
| 31400 | ADD no hyperactivity | attention deficit disorder without mention of hyperactivity |
| 31401 | ADD hyperactivity | attention deficit disorder with hyperactivity |
| 3272 | Organic sleep apnea | Organic sleep apnea |
| 32723 | obstructive sleep apnea (adult) (pediatric) | obstructive sleep apnea (adult) (pediatric) |
| 3320 | | Paralysis agitans |
| 3382 | Chronic pain | Chronic pain |
| 33829 | other chronic pain | other chronic pain |
| 340 | multiple sclerosis | multiple sclerosis |
| 3459 | unspecified epilepsy | unspecified epilepsy |
| 34590 | epilepsy: unspecified | epilepsy: unspecified without mention of intractable epilepsy |
| 3469 | unspecified migraine | unspecified migraine |
| 34690 | unspecified migraine | unspecified migraine without mention of intractable migraine without mention of status migrainosus |
| 3540 | carpal tunnel syndrome | carpal tunnel syndrome |
| 3559 | | Mononeuritis of unspecified site |
| 3569 | | Hereditary and idiopathic peripheral neuropathy: unspecified |
| 36250 | | Macular degeneration (senile)-unspecified |
| 3650 | Borderline glaucoma [glaucoma suspect] | Borderline glaucoma [glaucoma suspect] |
| 36500 | unspecified preglaucoma | unspecified preglaucoma |
| 3651 | Open-angle glaucoma | Open-angle glaucoma |
| 36511 | primary open angle glaucoma | primary open angle glaucoma |
| 3657 | | |
| 36570 | | |
| 3659 | unspecified glaucoma | unspecified glaucoma |
| 3661 | Senile cataract | Senile cataract |
| 36616 | senile cataract: nuclear sclerosis | senile cataract: nuclear sclerosis |
| 3669 | unspecified cataract | unspecified cataract |
| 3671 | myopia | myopia |

| | | |
|---|---|---|
| 3674 | | Presbyopia |
| 37230 | | Conjunctivitis unspecified |
| 3751 | Other disorders of lacrimal gland | Other disorders of lacrimal gland |
| 37515 | unspecified tear film insufficiency | unspecified tear film insufficiency |
| 3792 | Disorders of vitreous body | Disorders of vitreous body |
| 37921 | vitreous degeneration | vitreous degeneration |
| 38010 | | Infective otitis externa unspecified |
| 3804 | ear and mastoid process disease: impacted cerumen | diseases of the ear and mastoid process: impacted cerumen |
| 3818 | Other disorders of Eustachian tube | Other disorders of Eustachian tube |
| 38181 | dysfunction of eustachian tube | dysfunction of eustachian tube |
| 3829 | unspecified otitis media | unspecified otitis media |
| 3891 | Sensorineural hearing loss | Sensorineural hearing loss |
| 38910 | sensorineural hearing loss: unspecified | sensorineural hearing loss: unspecified |
| 3899 | unspecified hearing loss | unspecified hearing loss |
| 4011 | benign essential hypertension | benign essential hypertension |
| 4019 | essential hypt unspecify | essential hypertension: unspecified |
| 4039 | Hypertensive chronic kidney disease:unspecified | Hypertensive chronic kidney disease:unspecified |
| 40390 | unspecified hypertensive CKD | unspecified hypertensive chronic kidney disease with chronic kidney disease stage I through stage IV or unpescified |
| 4139 | | Other and unspecified angina pectoris |
| 4140 | Coronary atherosclerosis | Coronary atherosclerosis |
| 41400 | chronic ischemic heart d unspecify | chronic ischemic heart disease: unspecified type of vessel- native or graft |
| 41401 | coronary atherosclerosis of native coronary artery | coronary atherosclerosis of native coronary artery |
| 4241 | | Aortic valve disorders |
| 4254 | other primary cardiomyopathies | other primary cardiomyopathies |
| 4273 | Atrial fibrillation and flutter | Atrial fibrillation and flutter |
| 42731 | atrial fibrillation | atrial fibrillation |
| 4280 | congestive heart failure: unspecified | congestive heart failure: unspecified |
| 43491 | | Cerebral artery occlusion-unspecified with cerebral infarction |
| 4439 | | Peripheral vascular disease-unspecified |

| | | |
|---|---|---|
| 4619 | | Acute sinusitis unspecified |
| 462 | acute pharyngitis | acute pharyngitis |
| 4659 | unspecified acute respiratory infections | unspecified site of acute respiratory infections |
| 4660 | | Acute bronchitis |
| 470 | deviated nasal septum | deviated nasal septum |
| 4720 | chronic rhinitis | chronic rhinitis |
| 4739 | unspecified chronic sinusitis | unspecified chronic sinusitis |
| 4779 | unspecified upper respir tract D | unspecified cause of the upper respiratory tract disease |
| 4781 | Other diseases of nasal cavity and sinuses | Other diseases of nasal cavity and sinuses |
| 47819 | other disease of nasal cavity and sinuses | other disease of nasal cavity and sinuses |
| 486 | | Pneumonia organism unspecified |
| 490 | | Bronchitis not specified as acute or chronic |
| 4939 | unspecified asthma | unspecified asthma |
| 49390 | unspecified asthma | unspecified asthma |
| 496 | chronic airway obstruction: NEC | chronic airway obstruction: not elsewhere classified |
| 5308 | Other specified disorders of esophagus | Other specified disorders of esophagus |
| 53081 | esophageal reflux | esophageal reflux |
| 55090 | | Inguinal hernia without mention of obstruction or gangrene unilateral or unspecified |
| 56400 | | Constipation unspecified |
| 5859 | chronic kidney disease: unspecified | chronic kidney disease: unspecified |
| 5920 | calculus of kidney | calculus of kidney |
| 5939 | | Unspecified disorder of kidney and ureter |
| 5990 | urinary tract infection unspecified | urinary tract infection: stie not specified |
| 5997 | Hematuria | Hematuria |
| 59970 | hematuria: unspecified | hematuria: unspecified |
| 6000 | Hypertrophy (benign) of prostate | Hypertrophy (benign) of prostate |
| 60000 | hypertrophy (benign) of prostate without urinary obstruct-LUTS | hypertrophy (benign) of prostate without urinary obstruction and other lower urinary tract sympotoms (LUTS) |
| 60001 | hypertrophy (benign) of prostate with urinary obstruct-LUTS | hypertrophy (benign) of prostate with urinary obstruction and other lower urinary tract symptoms (LUTS) |
| 6078 | Other specified disorders of penis | Other specified disorders of penis |
| 60784 | impotence of organic origin | impotence of organic origin |
| 61610 | | Vaginitis and vulvovaginitis unspecified |

| | | |
|---|---|---|
| 6259 | unspecified symptom associated with female genital organs | unspecified symptom associated with female genital organs |
| 6272 | | Symptomatic menopausal or female climacteric states |
| 6929 | unspecified inflamma of skin and subcutaneous | unspecified cause of inflammatory conditions of skin and subcutaneous tissue |
| 6953 | rosacea | rosacea |
| 6961 | other psoriasis | other psoriasis |
| 7020 | actinic keratosis | actinic keratosis |
| 7021 | Seborrheic keratosis | Seborrheic keratosis |
| 70211 | inflamed seborrheic keratosis | inflamed seborrheic keratosis |
| 70219 | other seborrheic keratosis | other seborrheic keratosis |
| 7061 | other acne | other acne |
| 7062 | sebaceous cyst | sebaceous cyst |
| 7090 | Dyschromia | Dyschromia |
| 70909 | other dyschromia | other dyschromia |
| 7099 | unspecified disorder of skin and subcutaneous tissue | unspecified disorder of skin and subcutaneous tissue |
| 7140 | | Rheumatoid arthritis |
| 7159 | Osteoarthrosis: unspecified whether generalized or localized | Osteoarthrosis: unspecified whether generalized or localized |
| 71590 | osteoarthrosis: unspecified | osteoarthrosis: unspecified whether generalized or localized |
| 71596 | osteoarthrosis: lower leg | osteoarthrosis: lower leg |
| 7169 | unspecified arthropathy | unspecified arthropathy |
| 71690 | site unspecified arthropathy | site unspecified arthropathy |
| 7194 | Pain in joint | Pain in joint |
| 71941 | pain in joint in shoulder region | pain in joint in shoulder region |
| 71946 | pain in joint of lower leg | pain in joint of lower leg |
| 7231 | cervicalgia (pain in neck) | cervicalgia (pain in neck) |
| 7234 | | Brachia neuritis or radiculitis NOS |
| 7242 | lumbago | lumbago (other and unspecified disorders of back) |
| 7244 | thoracic or lumbosacral neuritis or radiculitis: unspecified | thoracic or lumbosacral neuritis or radiculitis: unspecified |
| 7245 | unspecified backache | unspecified backache |
| 7291 | myalgia and myositis: unspecified | myalgia and myositis: unspecified |

| | | |
|---|---|---|
| 7295 | pain in limb | pain in limb |
| 73300 | | Osteoporosis unspecified |
| 78039 | | Other convulsions |
| 7804 | dizziness and giddiness | dizziness and giddiness |
| 7805 | Sleep disturbances | Sleep disturbances |
| 78052 | unspecified insomnia | unspecified insomnia |
| 78057 | | Unspecified sleep apnea |
| 7807 | Malaise and fatigue | Malaise and fatigue |
| 78079 | other malaise and fatigue | other malaise and fatigue |
| 7810 | abnormal involuntary movements | abnormal involuntary movements |
| 7820 | disturbance of skin sensation | disturbance of skin sensation |
| 7821 | | Rash and other nonspecific skin eruption |
| 7823 | edema | edema |
| 7840 | headache | headache |
| 7851 | palpitations (awareness of heart beat) | palpitations (awareness of heart beat) |
| 7860 | Dyspnea and respiratory abnormalities | Dyspnea and respiratory abnormalities |
| 78605 | shortness of breath | shortness of breath |
| 78609 | other dyspnea and respiratory abnormalities | other dyspnea and respiratory abnormalities |
| 7862 | cough | cough |
| 7865 | Chest pain | Chest pain |
| 78650 | unspecified chest pain | unspecified chest pain |
| 7882 | Retention of urine | Retention of urine |
| 78820 | retention of urine: unspecified | retention of urine: unspecified |
| 7883 | Urinary incontinence | Urinary incontinence |
| 78830 | urinary incontinence: unspecified | urinary incontinence: unspecified |
| 7884 | Frequency of urination and polyuria | Frequency of urination and polyuria |
| 78841 | urinary frequency | urinary frequency |
| 7890 | Abdominal pain | Abdominal pain |
| 78900 | abdominal pain: unspecified site | abdominal pain: unspecified site |
| 7909 | Other nonspecific findings on examination of blood | Other nonspecific findings on examination of blood |
| 79093 | elevated PSA | elevated prostate specific antigen |
| 7962 | | Elevated blood pressure reading without diagnosis of hypertension |

| | | |
|---|---|---|
| 7999 | | Other unknown and unspecified cause |
| 8470 | | Sprains and strains of other and unspecified parts of back-Neck |
| V108 | Personal history of malignant neoplasm of other sites | Personal history of malignant neoplasm of other sites |
| V1083 | other malignant neoplasm of skin | other malignant neoplasm of skin |
| V1249 | | Other disorders of nervous system and sense organs |
| V202 | routine infant or child health check | routine infant or child health check |
| V221 | supervision of other normal pregnancy | supervision of other normal pregnancy |
| V238 | Other high-risk pregnancy | Other high-risk pregnancy |
| V2389 | other high-risk pregnancy | other high-risk pregnancy |
| V431 | Lens influence health | persons with a condition influencing their health status (Lens) |
| V456 | States following surgery of eye and adnexa | States following surgery of eye and adnexa |
| V4561 | cataract extraction status | cataract extraction status |
| V458 | Other postprocedural status | Other postprocedural status |
| V4589 | other postprocedural status | other postprocedural status |
| V508 | other elective surgery | other elective surgery for purposes other than remedying health states |
| V583 | | Attention to dressings and sutures |
| V586 | Long-term (current) drug use | Long-term (current) drug use |
| V5869 | long-term (current) use of other medications | long-term (current) use of other medications |
| V588 | Other specified procedures and aftercare | Other specified procedures and aftercare |
| V5883 | | Encounter for therapeutic drug monitoring |
| V5889 | other specified aftercare | other specified aftercare |
| V6549 | | Other specified counseling |
| V670 | Following surgery | Following surgery |
| V6700 | following surgery exam: unspecified | following surgery examination: unspecified |
| V6709 | following up examinations: other surgery | following up examinations: other surgery |
| V675 | Following other treatment | Following other treatment |
| V6759 | other follow up examination | other follow up examination |
| V679 | unspecified follow-up examination | unspecified follow-up examination |
| V700 | routine medical exam | routine general medical examination at a health care facility |
| V7189 | | Other specified suspected conditions |
| V720 | | Examination of eyes and vision |
| V723 | Gynecological examination | Gynecological examination |

| | | |
|---|---|---|
| V7231 | routine gynecological exam | routine gynecological examination |
| V7647 | | Special screening for malignant neoplasms-vagina |
| V99 | | |
| V990 | | |
| V997 | | |
