## Supplementary material for "Data-driven hypothesis generation among inexperienced clinical researchers: A comparison of secondary data analyses with visualization (VIADS) and other tools": training material (Supplemental Material 2),

VIADS

a **Visual Interactive Analysis** tool for filtering and summarizing large **Data Sets** coded with hierarchical terminologies

### VIADS study session-training

2021-12-02 @Clemson

VIADS

a **Visual Interactive Analysis** tool for filtering and summarizing large **Data Sets** coded with hierarchical terminologies

### Background

- Visualization
  - Versus excel files
  - Hierarchical relationships
- Data sets coded by hierarchical ***terminologies*** in health care
  - ICD9 (monohierarchic)
  - ICD10 (monohierarchic)
  - MeSH (polyhierarchic)
- However

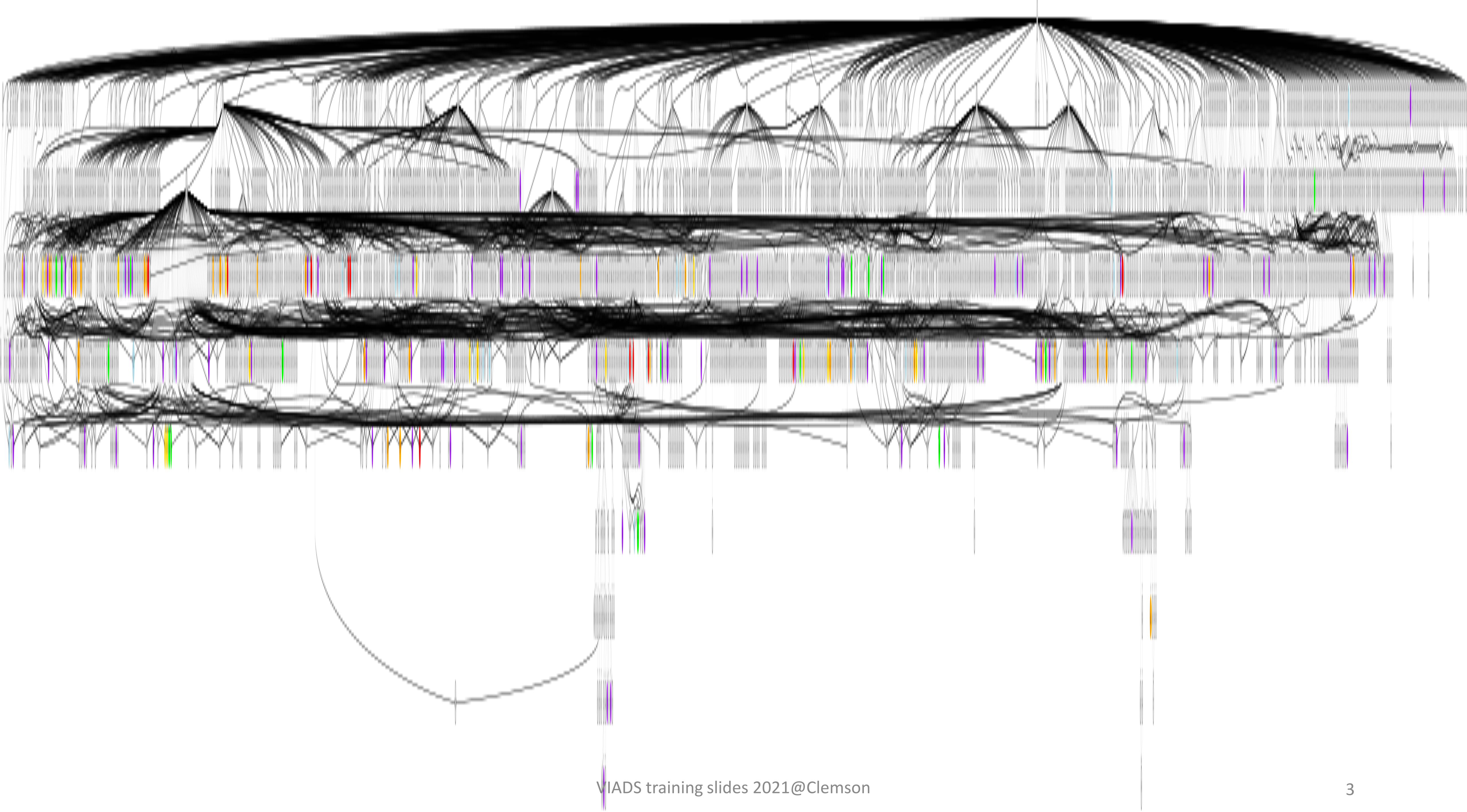

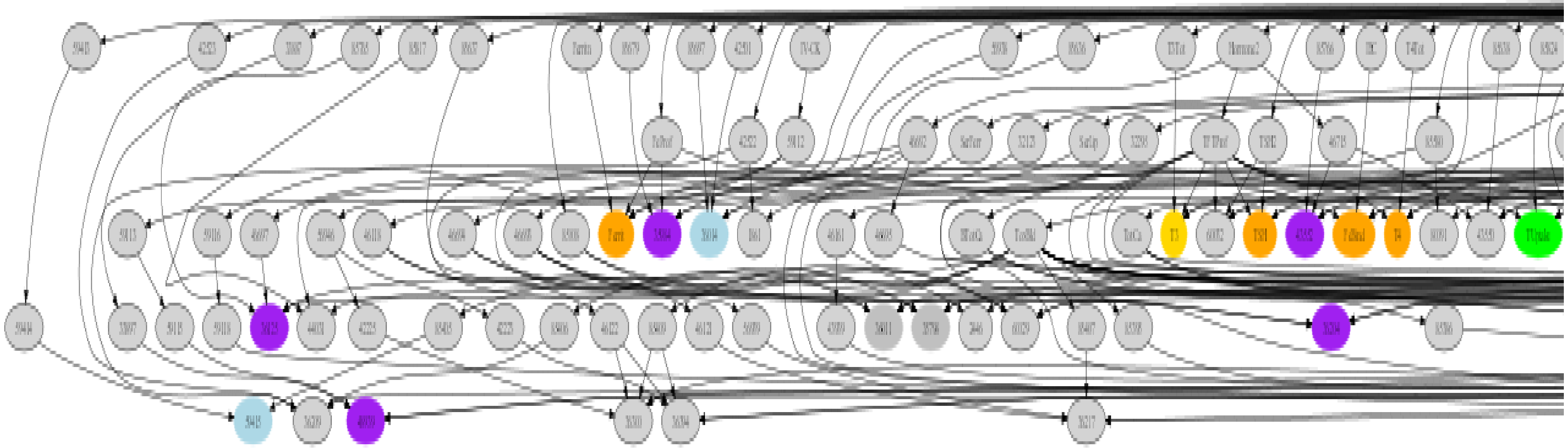

VIADS

a **Visual Interactive Analysis** tool for filtering and summarizing large **Data Sets** coded with hierarchical terminologies

### Background (Cont.)

- Algorithms development
  - **Reduce** size
  - **Simplify** complexity
  - **Analyze** large data set
    - Summarize
    - Compare & highlight

  - By using hierarchical **terminologies** & **frequencies**

### Filter --- **node counts (NC)**

If NC >= $T_{nc}$, then display node n.

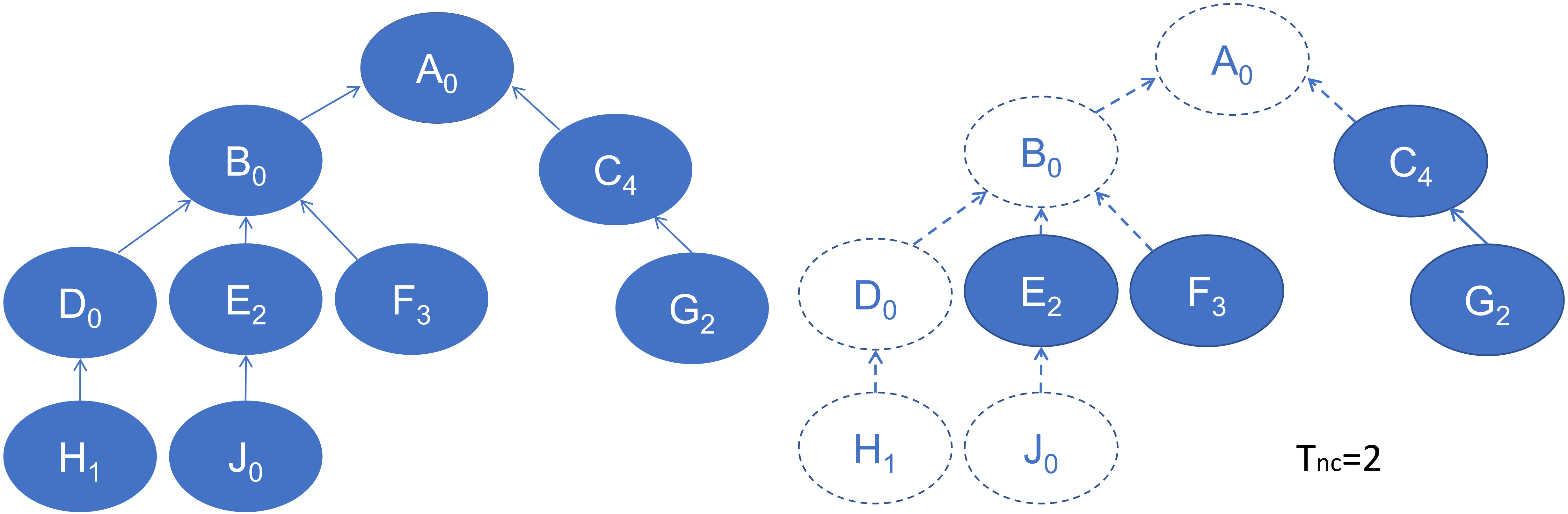

### Filter --- class counts (CC)

- $CC_n = NC_{dn1} + NC_{dn2} + NC_{dn3} \ldots NC_{dnk}$

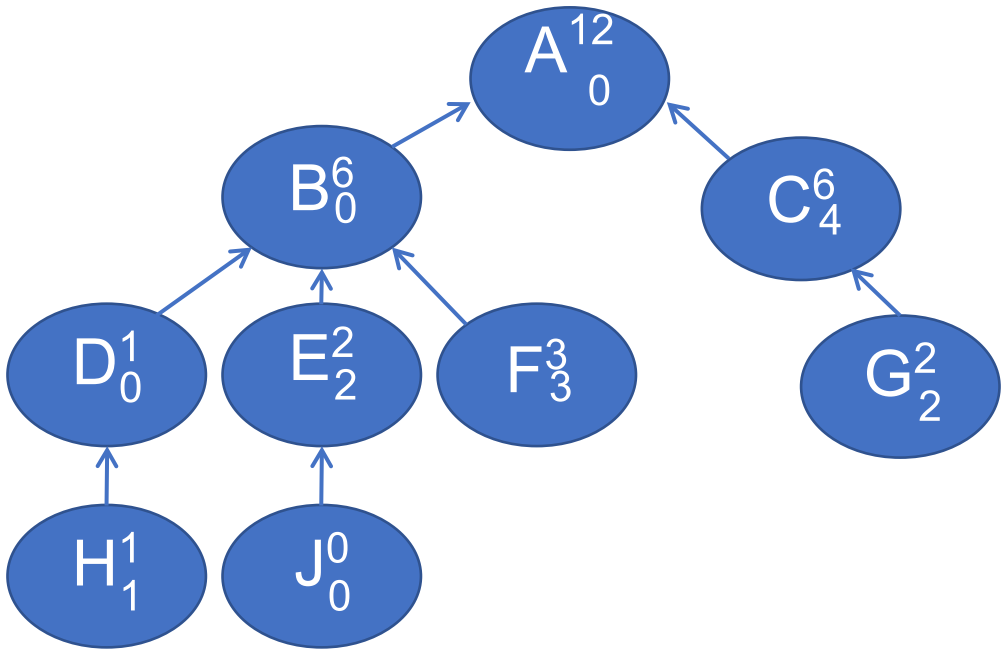


**Table 1** Data table

| Node | Node count | Class count |
|---|---|---|
| A | 0 | 12 |
| B | 0 | 6 |
| C | 4 | 6 |
| D | 0 | 1 |
| E | 2 | 2 |
| F | 3 | 3 |
| G | 2 | 2 |
| H | 1 | 1 |
| J | 0 | 0 |

### Filtering --- class counts

If $CC_n$ >=$T_{cc}$, then display n and *all the ancestors* of n in the terminological structure.

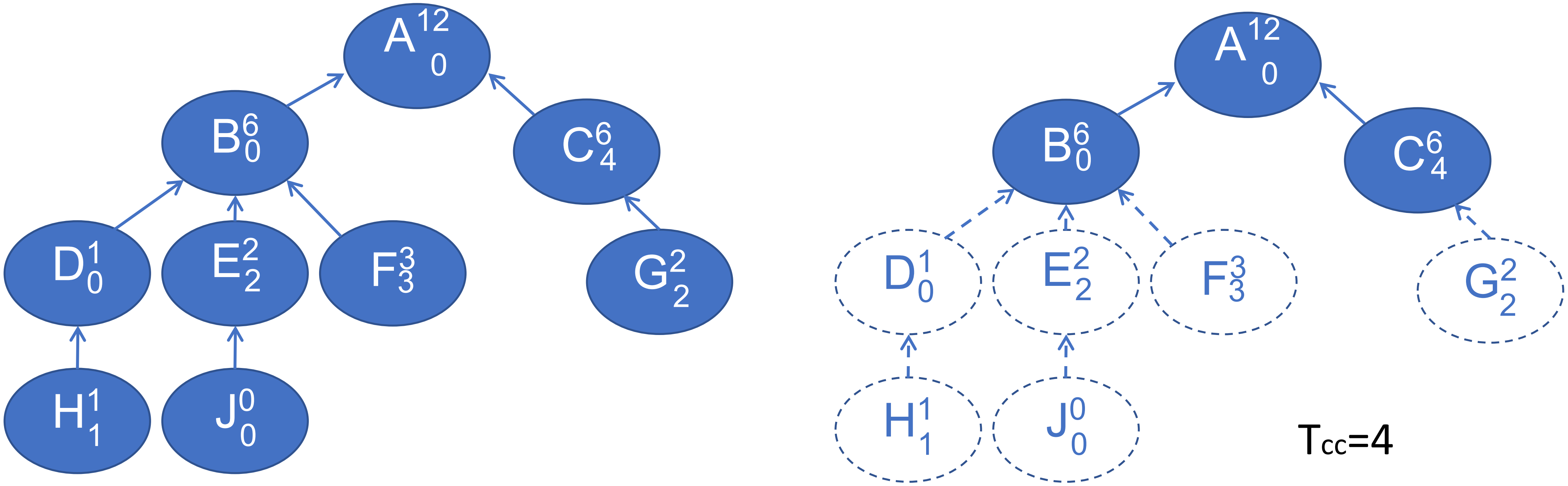

### Filter --- **ratio values (RV)**

- $RV_n = CC_n / CC_p$

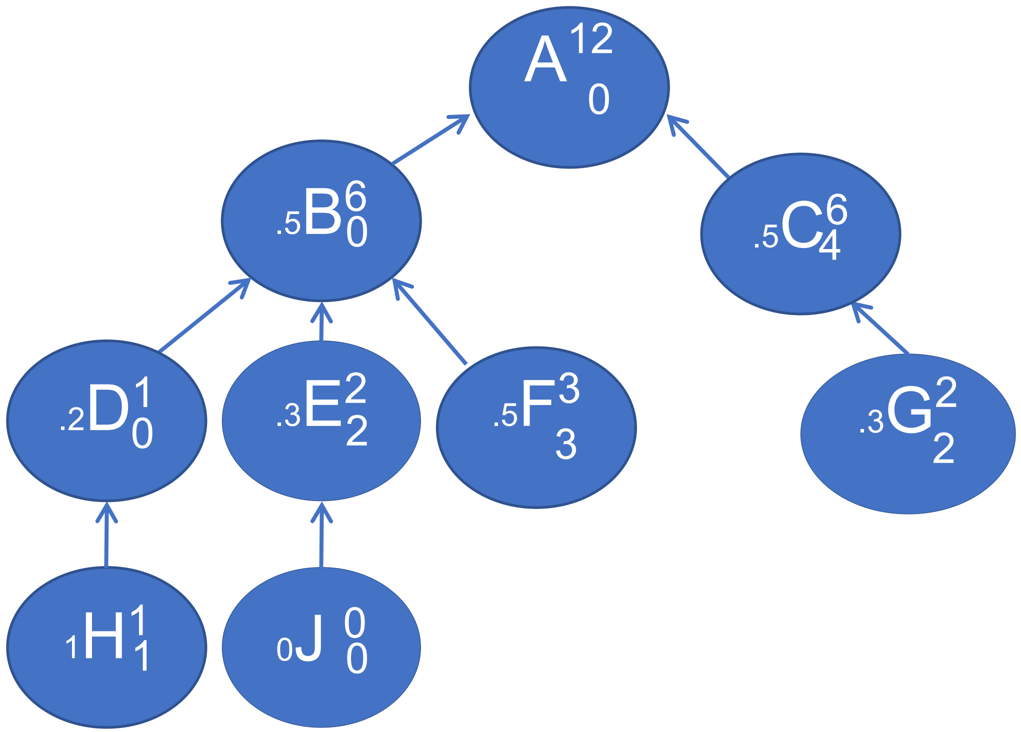


**Table 1** Calculated ratio table

| Node | CC | ratio |
|---|---|---|
| B | 6 | 0.5 |
| C | 6 | 0.5 |
| D | 6 | 0.2 |
| E | 2 | 0.3 |
| F | 3 | 0.5 |
| G | 2 | 0.3 |
| H | 1 | 1 |
| J | 0 | 0 |

### Filter --- **ratio values**

If $RV_n >= T_r$, then display n and all the ancestors of n in the terminological structure.

$RV_n = CC_n / CC_p$

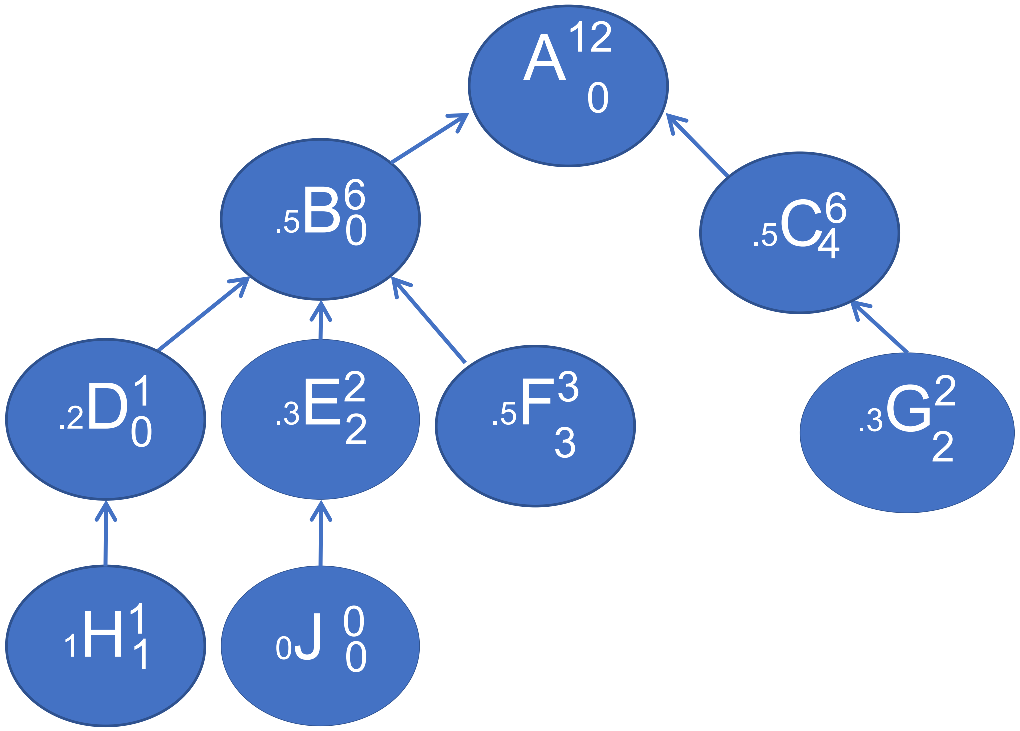


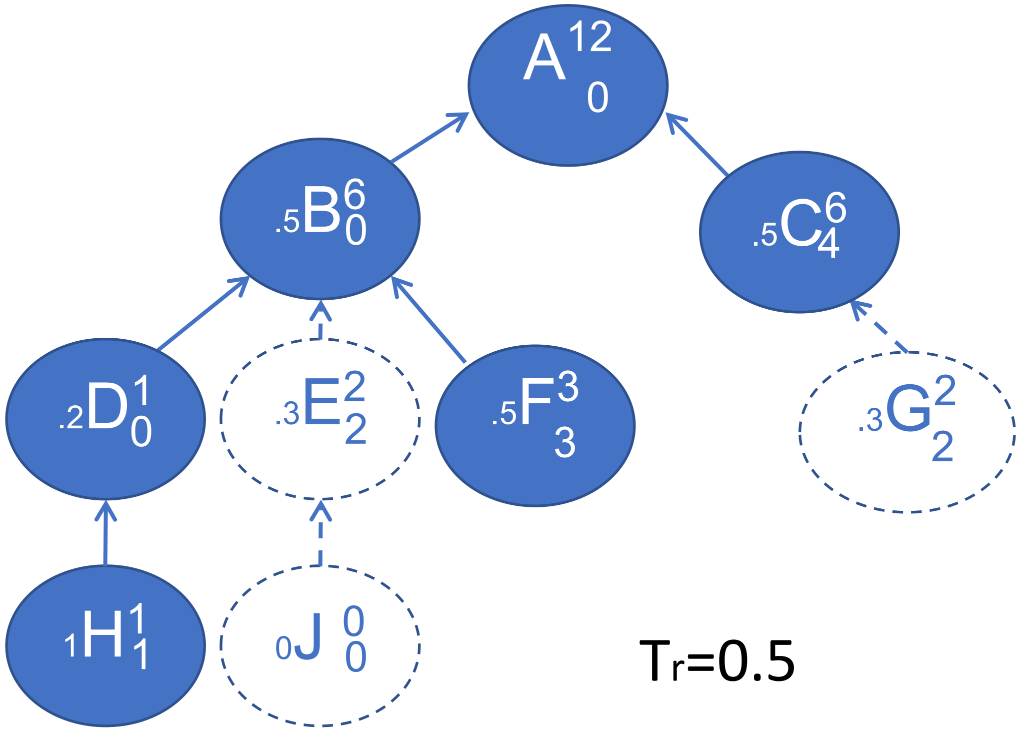

### Comparison of similar data sets(diff)

- Comparison of two similar data sets
  - Two decades, two similar medications (Actos, Avandia), before and after a procedure …

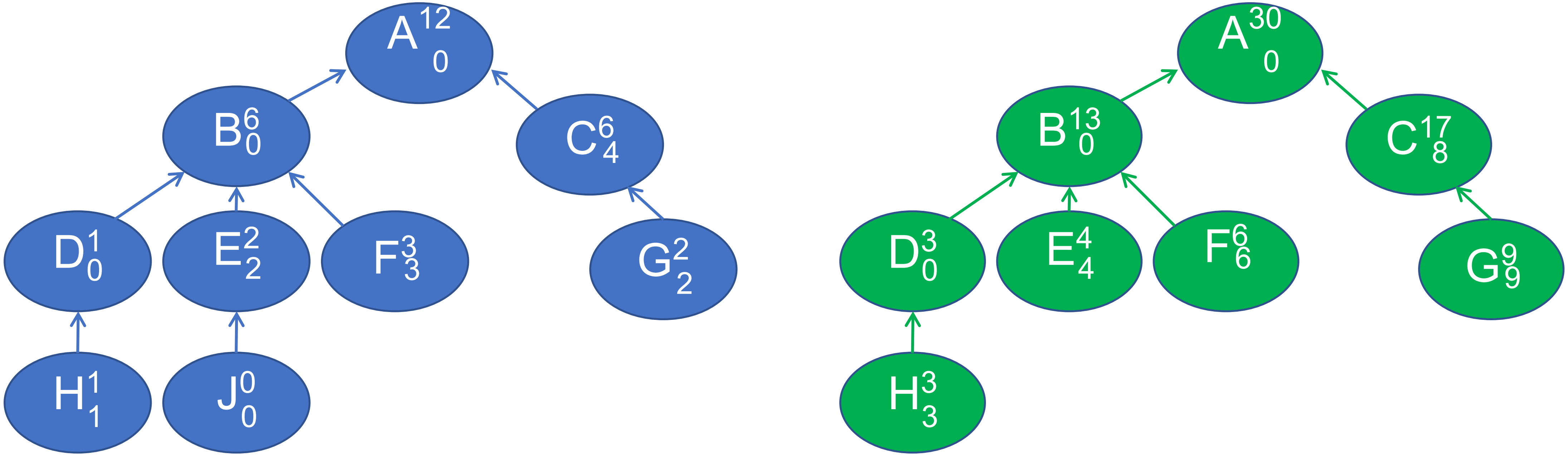

### Filter --- p values for diff data sets (prop.test)

If $P_n >= T_p$, then display n and all the ancestors of n in the terminological structure.

$$CCP(term) = CC(term) / \sum_{k=1}^{n} CC(term)\ k$$

| Node | CC in data set 1 | CC in data set 2 | P value |
|---|---|---|---|
| A | 12 | 30 | 1 |
| B | 6 (6/12) | 13 (13/30) | 0.0065 |
| C | 6 (6/12) | 17 (17/30) | 0.0219 |
| D | 1 (1/12) | 3 (3/30) | 1 |
| E | 2 (2/12) | 4 (4/30) | 0.8521 |
| F | 3 (3/12) | 6 (6/30) | 0.6062 |
| G | 2 (2/12) | 9 (9/30) | 1 |
| H | 1 (2/12) | 3 (3/30) | 1 |

### CC+RV --- thresholds setting

- 3D plot (scatter3dplot, R)- highlight points
  - CC, RV, the number of nodes in filtered graphs
- Regression plane
  - Minimizes the total of the squared distances: the observed values → the closest point on the regression plane
  - The least squared distance (residual, R)
- Select data points
  - **90-120** + **closest to the plane**

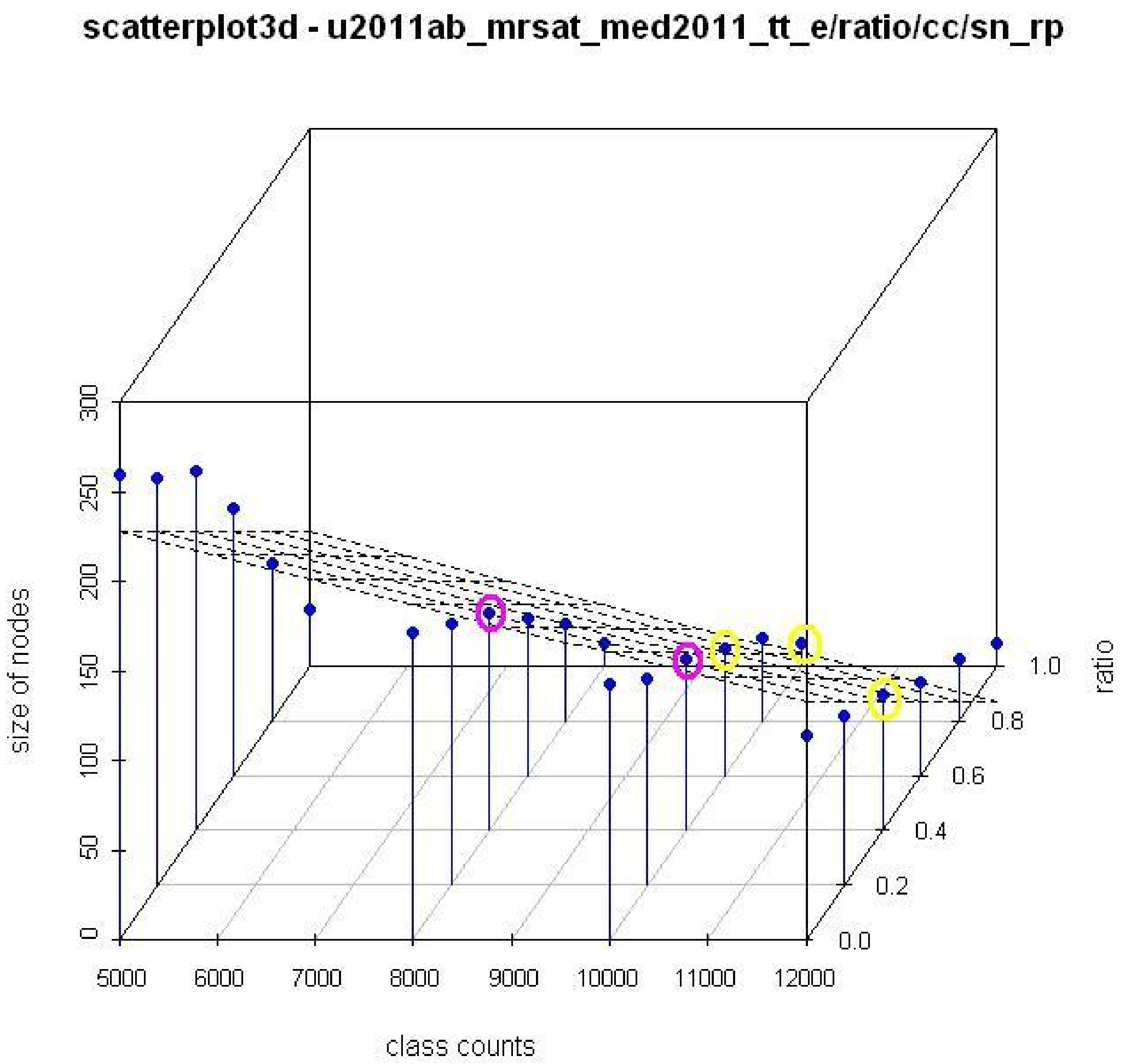


3D plot of class count thresholds, ratio value thresholds and the number of nodes in filtered graphs for MeSH terms in 2011 with the regression plane.

### Purposes of using VIADS

- Facilitate to understand the data sets
- See the bigger picture, aggregated effects
- Highlight the differences
- By trimming down the relatively unimportant parts

- To generate *new research ideas*

**Diseases and Injuries Tabular Index**

1. INFECTIOUS AND PARASITIC DISEASES (001-139)
2. NEOPLASMS (140-239)
3. ENDOCRINE, NUTRITIONAL AND METABOLIC DISEASES, AND IMMUNITY DISORDERS (240-279)
4. DISEASES OF THE BLOOD AND BLOOD-FORMING ORGANS (280-289)
5. MENTAL DISORDERS (290-319)
6. DISEASES OF THE NERVOUS SYSTEM AND SENSE ORGANS (320-389)
7. DISEASES OF THE CIRCULATORY SYSTEM (390-459)
8. DISEASES OF THE RESPIRATORY SYSTEM (460-519)
9. DISEASES OF THE DIGESTIVE SYSTEM (520-579)
10. DISEASES OF THE GENITOURINARY SYSTEM (580-629)
11. COMPLICATIONS OF PREGNANCY, CHILDBIRTH, AND THE PUERPERIUM (630-679)
12. DISEASES OF THE SKIN AND SUBCUTANEOUS TISSUE (680-709)
13. DISEASES OF THE MUSCULOSKELETAL SYSTEM AND CONNECTIVE TISSUE (710-739)
14. CONGENITAL ANOMALIES (740-759)
15. CERTAIN CONDITIONS ORIGINATING IN THE PERINATAL PERIOD (760-779)
16. SYMPTOMS, SIGNS, AND ILL-DEFINED CONDITIONS (780-799)
17. INJURY AND POISONING (800-999)

SUPPLEMENTARY CLASSIFICATION OF FACTORS INFLUENCING HEALTH STATUS AND CONTACT WITH HEALTH SERVICES (V01-V89)

SUPPLEMENTARY CLASSIFICATION OF EXTERNAL CAUSES OF INJURY AND POISONING (E800-E999)

#### 1. INFECTIOUS AND PARASITIC DISEASES (001-139)

##### TUBERCULOSIS (010-018)

Includes:

infection by Mycobacterium tuberculosis (human) (bovine)

Excludes:

*congenital tuberculosis (771.2)*
*late effects of tuberculosis (137.0-137.4)*

The following fifth-digit subclassification is for use with categories 010-018:

0 unspecified
1 bacteriological or histological examination not done
2 bacteriological or histological examination unknown (at present)
3 tubercle bacilli found (in sputum) by microscopy
4 tubercle bacilli not found (in sputum) by microscopy, but found by bacterial culture
5 tubercle bacilli not found by bacteriological examination, but tuberculosis confirmed histologically
6 tubercle bacilli not found by bacteriological or histological examination, but tuberculosis confirmed by other methods [inoculation of animals]

- **010 Primary tuberculous infection**
  Requires fifth digit. See beginning of section 010-018 for codes and definitions.
- **011 Pulmonary tuberculosis**
  Requires fifth digit. See beginning of section 010-018 or codes and definitions.
  Use additional code to identify any associated silicosis (502)
- **012 Other respiratory tuberculosis**
  Requires fifth digit. See beginning of section 010-018 for codes and defini
  Excludes:
  *respiratory tuberculosis, unspecified (011.9)*
- **013 Tuberculosis of meninges and central nervous system**
  Requires fifth digit. See beginning of section 010-018 for codes and defini

**TUBERCULOSIS (010-018)**

**010 Primary tuberculous infection**
Requires fifth digit. See beginning of section 010-018 for codes and definitions.

**010.0 Primary tuberculous infection**
[0-6]
Excludes:
*nonspecific reaction to tuberculin skin test without active tuberculosis (795.5)*
*positive PPD (795.5)*
*positive tuberculin skin test without active tuberculosis (795.5)*

**010.1 Tuberculous pleurisy in primary progressive tuberculosis**
[0-6]

**010.8 Other primary progressive tuberculosis**
[0-6]
Excludes:
*tuberculous erythema nodosum (017.1)*

**010.9 Primary tuberculous infection, unspecified**
[0-6]
