## Supplementary material for "Data-driven hypothesis generation among inexperienced clinical researchers: A comparison of secondary data analyses with visualization (VIADS) and other tools": study scripts (Supplemental Material 3),

### Study session script for groups 3 and 4 (with VIADS)

Updated on 2021-10-05

This document explains the preparation before the study session and activities during and after the study session for the research participant, i.e., you. The study session refers to the part during which both audio and screen activities will be recorded. This document includes the following six sections: prior to the study session, checklist, VIADS training session, during the study session, after the study session, and the dimensions a hypothesis can consider. Please read through the document and understand the study; bring up any questions you may have before or during the study session.

#### Prior to the study session

This section details the files you will receive and the preparation you should do before the study session. You should have the following tools or infrastructure ready for use:

1. Operational system: Windows 8 or 10
2. Web browser: Google Chrome
3. Stable internet connection
4. The corresponding **informed consent form** sent to you
5. The **study session script** sent to you
6. The data sets that will be used to generate hypotheses or research ideas in the study session: two ICD9 data sets (ICD9 codes and frequencies) and one file that includes the full names of the ICD9 codes used.
7. You should also receive a test(demonstration) data set, which will be used during the training session.
8. The researcher should answer any questions that you may have about the script. During the study session, you will use the given data sets to generate hypotheses, and the researcher will observe and record the process. The conversations, i.e., audio and screen "think aloud" activities, will be recorded. The researcher will clarify any questions you may have. The researcher may
   a. Ask follow up inquiry questions
   b. Ask heuristic questions
   c. Provide suggestions about possible options in formulating a hypothesis
   d. Clarify any questions you may have
9. With your help, the researcher will set up the study session date/time (scheduling)
   a. Date/time of training session (Approximately 1 hour)
   b. Date/time of study session (Approximately 2 hours)
10. You can select the gift card options in discussion with the RA, Brooke Dragi.
    a. Type of gift card
    b. By email or by mail, a mailing address is needed if a physical card is selected.
11. You should have access to a quiet space during the study session and a pen and blank paper for you to use
12. You practice think-aloud protocol
    a. To verbally "work through" and articulate what you are doing while doing it
13. You test Internet connection and audio

14. You test Webex software

#### Your checklist for the study session

1. Software: WebEx, Google Chrome
2. Equipment: microphone, Internet connections
3. Data sets
4. Blank papers and pen/pencil
5. Confirm study session day/time
6. Please bring in any questions you may have to the study session.

#### VIADS training session

1. The researcher will go through the study flow in VIADS by using the test (demonstration) data set with you
    a. Log in as a guest user
    b. Upload the training data set
    c. Validate the data set
    d. Select at least two to three algorithms (e.g., top NC, Class counts, CC + ratio)
        i. Top NC
        ii. CC
        iii. CC + ratio
    e. Tune thresholds and preview corresponding results
        i. Top 25, 40, 50
        ii. CC
        iii. CC and ratio (CC+ 0.8, 0.9, 0.95)
        iv. Calculate the number of nodes
        v. Results preview (2D, 3D)
        vi. Generate from data
    f. Results interpretation
        i. Possible hypothesis
        ii. Refer to hypothesis generation consideration dimensions to formulate a hypothesis (or more than one)
    g. Graph manipulation
        i. Zoom in/out
        ii. Option area appears/disappear
        iii. Different horizontal distances
    h. Graph interpretations and visualization features
        i. Possible hypothesis
        ii. Refer to hypothesis generation consideration dimensions to formulate a hypothesis (or more than one)
        iii. Refine hypothesis
    i. Save the results (PNG, SVG files, data, pay attention to download path)
    j. Pause intermittently and give the participant time to reflect
2. The researcher guides the process (point 1 in this section) first
3. Then you will go through the process by using the VIADS without external help

    a. From uploading the data set until saving the results (graphs and data file)
4. While you use VIADS, you practice think-aloud protocol at the same time
    a. You speak out the annotations when you do the analysis
    b. There might be conversations between you and the researcher
5. Screen activities and audio will be recorded through the one-hour training session
6. The researcher will double-check if you have any questions or concerns

#### During the study session

1. Greetings
2. The researcher will restate the study flow, what will be recorded, and confirm consent verbally
3. The researcher states the plan of the study session during the two-hour session, i.e., you use VIADS to analyze the data sets to generate hypotheses
    a. The following are possible scenarios to generate hypotheses
        i. based on the analytic results
        ii. derived from the analysis
        iii. provoked new research ideas by the analysis (not necessarily relevant, most likely irrelevant)
    b. You can refer to hypothesis generation consideration dimensions to formulate or refine a hypothesis (or more than one)
4. You will use VIADS to do the following steps during the study session:
    a. Log in as a guest user
    b. Upload and validate the study data set, one or both
    c. Select at least two algorithms to analyze the data set (e.g., top NC, CC, CC+ratio)
    d. Tune thresholds under each algorithm to generate graphs
    e. Generate hypothesis and use think-aloud protocol during the graph generation and the result interpretation processes
        i. Directly based on the results
        ii. Derived from the analytic results
        iii. Provoked by the process
    f. Save results if needed
5. You can generate one or more hypotheses. The hypothesis generation task is deemed as complete when you determine so, or the study session reaches two hours.
6. After you complete the hypothesis generation task, the following questions will be asked by the researcher:
    a. What activities/events (e.g., read papers/books, discuss with colleagues/students/family, presentation Q & A, daily work, witness something completely irrelevant) provoked the new research ideas in the past?
    b. How do you capture research ideas in the first place usually?
    c. Do you describe yourself as a creative person versus someone who follows the instructions carefully in general? I.e., a creative person may always try to find new ways to do things or to look at things. (Likert scale 1 to 5, the lowest to the highest)
    d. Do you know if others, e.g., your family, close friends, colleagues, perceive you as a creative person? (yes, no, not sure)
    e. What will facilitate the generation of new research ideas in your view?

#### Study session script for group 3 and 4 (with VIADS)

7. Screen activities and audio will be recorded through the two-hour study session
8. All results will be anonymized, categorized, and aggregated during analyzing and publishing

#### After the study session

1. You will be asked to complete the VIADS System Usability Scale (SUS) questionnaire
2. You will be asked to complete the follow-up survey
3. Compensation will be determined based on the actual time spent to calculate the, to obtain and distribute gift cards accordingly.
4. Brook will log the details about gift cards in the log. The following information will be recorded:
    a. Participant number (ID)
    b. Hours spent, i.e., training session, study session, time to complete surveys
    c. Training session and study session date/time
    d. Gift card amount and the last four digits of the gift cards
    e. Gift card type
    f. Gift card distribution date/type
    g. Gift card receiving confirmation

#### Hypothesis generation consideration dimensions:

- Basic, applied, translational research
- Observational or experimental study
- Descriptive or analytic research
- Comparison, association, or causal study
- Retrospective or prospective research
- Longitudinal or cross-sectional research
- Qualitative or quantitative research
- Single variable or multiple variables
- Variables: dependent, independent, moderator, control, and intervening
- Focused population
- One-tailed or two-tailed
- Investigational intervention
- Outcomes of interest
