## Supplementary material for "Data-driven hypothesis generation among inexperienced clinical researchers: A comparison of secondary data analyses with visualization (VIADS) and other tools": follow-up surveys (Supplemental Material 4 & 5),

### VIADS_Participant_follow_up_survey

Start of Block: Default Question Block

Q1 Your job title:

________________________________________________________________

Q2 The type of organization that you affiliate with (check all that apply):

▢ Academic institution (1)

▢ Academic medical center (2)

▢ Hospital (3)

▢ Practice (4)

▢ Non-profit organization (5)

▢ Other, please specify (6) ________________________________________________

Q3 How often do you use computers?

- ○ Daily, < 2 hours (1)
- ○ Daily, 2-4 hours (2)
- ○ Daily, > 4 hours (3)
- ○ Weekly (4)
- ○ Monthly (5)

---

Q4
How many years of experience do you have in clinical research: hypothesis generation?

- ○ (1)
- ○ > 2 years and < 5 years (2)
- ○ >= 5 years and < 10 years (3)
- ○ >= 10 years (4)

---

Q5 How many years of experience do you have in clinical research: study design?

- ○ (1)
- ○ > 2 years and < 5 years (2)
- ○ >= 5 years and < 10 years (3)
- ○ >= 10 years (4)

---

Q6
How many years of experience do you have in data analysis during clinical research?

- (1)
- > 2 years and < 5 years (2)
- >= 5 years and < 10 years (3)
- >= 10 years (4)

---

Q7
What type of role do you mainly play in clinical research during hypothesis generation?

- Leading role (1)
- Participant role (2)
- Other, please specify (3) ________________________________________________

---

Q8 What type of role do you mainly play in clinical research during study design?

- Leading role (1)
- Participant role (2)
- Other, please specify (3) ________________________________________________

---

Q9
What data analysis tools do you use more often? Please check all that apply

- ☐ MS Excel (1)
- ☐ R (2)
- ☐ SAS (3)
- ☐ SPSS (4)
- ☐ Other, please specify (5) ________________

---

Q10 If you were provided with more detailed information about research design (such as focused population) during your hypothesis generation process, do you think the information would be helpful in formulating your hypothesis overall?

- ○ Yes (1)
- ○ Maybe, please explain (2) ________________
- ○ No, please explain (4) ________________

---

Q11
In a scale of 0 (the least helpful) to 10 (the most helpful), how helpful do you think it would be to use each of the following dimensions in formulating your hypothesis:

0 1 2 3 4 5 6 7 8 9 10

| |
|---|
| Basic, applied, translational research () |
| Observational or experimental study () |
| Descriptive or analytic research () |
| Comparison, association, or causal study () |
| Retrospective or prospective research () |
| Longitudinal or cross-sectional research () |
| Qualitative or quantitative research () |
| Single variable or multiple variables () |
| Variables: dependent, independent, moderator, control, and intervening () |
| Focused population () |
| One-tailed or two-tailed () |
| Investigational intervention () |
| Outcomes of interest () |

Q12
Can you give examples of helpful instructions in formulating a hypothesis?

________________________________________________________________

________________________________________________________________

________________________________________________________________

________________________________________________________________

________________________________________________________________

Q13
Do you have any additional comments or suggestions about how to facilitate hypothesis generation?

________________________________________________________________

________________________________________________________________

________________________________________________________________

________________________________________________________________

________________________________________________________________

**End of Block: Default Question Block**
