## Supplementary material for "Data-driven hypothesis generation among inexperienced clinical researchers: A comparison of secondary data analyses with visualization (VIADS) and other tools": follow-up surveys (Supplemental Material 4 & 5),

### VIADS system usability scale (SUS) and utility questionnaire

Start of Block: Default Question Block

Q1 I would like to use VIADS frequently.

- Strongly disagree (1)
- Disagree (2)
- Neutral (3)
- Agree (4)
- Strongly agree (5)

*Display This Question:*

*If I would like to use VIADS frequently. = Strongly disagree*

*Or I would like to use VIADS frequently. = Disagree*

Q2 Can you specify the reasons for your answer (i.e., why will not you use VIADS frequently)?

________________________________________________________________

________________________________________________________________

________________________________________________________________

________________________________________________________________

________________________________________________________________

Q3 I find VIADS unnecessarily complex.

- ◯ Strongly disagree (1)
- ◯ Disagree (2)
- ◯ Neutral (3)
- ◯ Agree (4)
- ◯ Strongly agree (5)

---

*Display This Question:*

*If I find VIADS unnecessarily complex. = Strongly agree*

*Or I find VIADS unnecessarily complex. = Agree*

Q4 Please specify what you would do to make VIADS simpler. Please list three things you would change in VIADS if possible.

________________________________________________________________

________________________________________________________________

________________________________________________________________

________________________________________________________________

________________________________________________________________

---

Q5 I think VIADS is easy to use.

- ◯ Strongly disagree (1)
- ◯ Disagree (2)
- ◯ Neutral (3)
- ◯ Agree (4)
- ◯ Strongly agree (5)

*Display This Question:*

*If I think VIADS is easy to use. = Strongly disagree*

*Or I think VIADS is easy to use. = Disagree*

Q6 Can you please give an example about VIADS is not easy to use?

________________________________________________________________

________________________________________________________________

________________________________________________________________

________________________________________________________________

________________________________________________________________

Q7
I would need technical support in order to use VIADS.

- ◯ Strongly disagree (1)
- ◯ Disagree (2)
- ◯ Neutral (3)
- ◯ Agree (4)
- ◯ Strongly agree (5)

*Display This Question:*

*If I would need technical support in order to use VIADS. = Strongly agree*

*Or I would need technical support in order to use VIADS. = Agree*

Q8 Can you please specify the support (such as navigation, definitions of the terms used in VIADS interfaces, mechanism of VIADS behind the scenes ) you need?

________________________________________________________________

________________________________________________________________

Q9 I find the various functions in VIADS are well integrated with each other.

- Strongly disagree (1)
- Disagree (2)
- Neutral (3)
- Agree (4)
- Strongly agree (5)

*Display This Question:*

*If I find the various functions in VIADS are well integrated with each other. = Strongly disagree*

*Or I find the various functions in VIADS are well integrated with each other. = Disagree*

Q10
Can you please give an example to elaborate your answer? So we can try to look for ways to improve. Please feel free to add suggestions to make them better integrated, ideally.

Q11 I think there are too many inconsistencies in VIADS.

- ◯ Strongly disagree (1)
- ◯ Disagree (2)
- ◯ Neutral (3)
- ◯ Agree (4)
- ◯ Strongly agree (5)

---

*Display This Question:*

*If I think there are too many inconsistencies in VIADS. = Strongly agree*

*Or I think there are too many inconsistencies in VIADS. = Agree*

Q12 Can you please give examples of inconsistencies in VIADS?

________________________________________________________________

________________________________________________________________

________________________________________________________________

________________________________________________________________

________________________________________________________________

---

Q13 I imagine that most people can learn to use VIADS quickly.

- ◯ Strongly disagree (1)
- ◯ Disagree (2)
- ◯ Neutral (3)
- ◯ Agree (4)
- ◯ Strongly agree (5)

---

*Display This Question:*

*If I imagine that most people can learn to use VIADS quickly. = Strongly disagree*

*Or I imagine that most people can learn to use VIADS quickly. = Disagree*

Q14 Can you please explain why you think most people cannot learn to use VIADS quickly?

________________________________________________________________

________________________________________________________________

________________________________________________________________

________________________________________________________________

________________________________________________________________

Q15 I find VIADS cumbersome to use.

- ◯ Strongly disagree (1)
- ◯ Disagree (2)
- ◯ Neutral (3)
- ◯ Agree (4)
- ◯ Strongly agree (5)

*Display This Question:*

*If I find VIADS cumbersome to use. = Strongly agree*

*Or I find VIADS cumbersome to use. = Agree*

Q16 Can you please give an example that VIADS is cumbersome to use?

________________________________________________________________

________________________________________________________________

________________________________________________________________

________________________________________________________________

Q17 I feel confident using VIADS.

- Strongly disagree (1)
- Disagree (2)
- Neutral (3)
- Agree (4)
- Strongly agree (5)

*Display This Question:*

*If I feel confident using VIADS. = Strongly disagree*

*Or I feel confident using VIADS. = Disagree*

Q18 Can you please explain why you are not confident using VIADS?

Q19 I need to learn more before I can use VIADS.

- ◯ Strongly disagree (1)
- ◯ Disagree (2)
- ◯ Neutral (3)
- ◯ Agree (4)
- ◯ Strongly agree (5)

---

*Display This Question:*

*If I need to learn more before I can use VIADS. = Strongly agree*

*Or I need to learn more before I can use VIADS. = Agree*

Q20 Can you please specify what you need to learn before you can use VIADS? An example will be very helpful.

________________________________________________________________

________________________________________________________________

________________________________________________________________

________________________________________________________________

________________________________________________________________

---

Q21 Does VIADS provide new perspectives or measurements for understanding the data set?

- ◯ Yes (1)
- ◯ Maybe (2)
- ◯ No (3)

---

*Display This Question:*

*If Does VIADS provide new perspectives or measurements for understanding the data set? = Yes*

*Or Does VIADS provide new perspectives or measurements for understanding the data set? = Maybe*

Q22 Can you specify your answer to provide more details please?

________________________________________________________________

________________________________________________________________

________________________________________________________________

________________________________________________________________

________________________________________________________________

Q23 Do you think that VIADS can facilitate the interpretation of the data set?

- Yes (1)
- Maybe (2)
- No (3)

*Display This Question:*

*If Do you think that VIADS can facilitate the interpretation of the data set? = Yes*

*Or Do you think that VIADS can facilitate the interpretation of the data set? = Maybe*

Q24 Can you specify your answer to provide more details please?

________________________________________________________________

________________________________________________________________

________________________________________________________________

________________________________________________________________

________________________________________________________________

Q25 Do you think that VIADS can facilitate your decision making in generating hypothesis?

- Yes (1)
- Maybe (2)
- No (3)

---

Q26 Can you specify your answer to provide more details please?

________________________________________________________________

________________________________________________________________

________________________________________________________________

________________________________________________________________

________________________________________________________________

---

Q28 Do you think that VIADS can facilitate your capability to present the data set?

- Yes (1)
- Maybe (2)
- No (3)

---

*Display This Question:*

*If Do you think that VIADS can facilitate your capability to present the data set? = Yes*

*Or Do you think that VIADS can facilitate your capability to present the data set? = Maybe*

Q29 Can you specify your answer to provide more details please?

________________________________________________________________

________________________________________________________________

________________________________________________________________

________________________________________________________________

________________________________________________________________

Q30 Do you think that VIADS can be used in other aspects of research questions in addition to facilitating understanding of data, interpretation of data, presentation of data, decisions on hypothesis generation?

- Yes (1)
- Maybe (2)
- No (3)

*Display This Question:*

*If Do you think that VIADS can be used in other aspects of research questions in addition to facilit... = Yes*

*Or Do you think that VIADS can be used in other aspects of research questions in addition to facilit... = Maybe*

Q31 Can you specify your answer to provide more details please?

________________________________________________________________

________________________________________________________________

________________________________________________________________

________________________________________________________________

________________________________________________________________

Q32 Do you think that VIADS is a useful tool to facilitate research activities in general?

- Yes (1)
- Maybe (2)
- No (3)

Q33 Do you have any additional suggestions or comments about VIADS?

________________________________________________________________

________________________________________________________________

________________________________________________________________

________________________________________________________________

________________________________________________________________

**End of Block: Default Question Block**
