## Supplementary material for "Data-driven hypothesis generation among inexperienced clinical researchers: A comparison of secondary data analyses with visualization (VIADS) and other tools": hypothesis quality evaluation instrument (Supplemental Material 6)

VIADS Expert feedback on hypothesis evaluation matrix

### 10-item comprehensive version of clinical hypotheses evaluation

. This is a comprehensive evaluation of the following hypothesis:

To compare different states in the USA if the incidence of 5859 (ICD9 code: chronic kidney diseases, unspecified) is correlated to kidney transplantation surgeries. If there are differences, what causes these differences? Donors? Surgeons? Or other reasons.

*Q1.1.* The following metrics are intended to evaluate the **validity** (the hypothesis seems logically well-founded and likely corresponds accurately to the real world, existing sciences, or clinical experiences without being fundamentally against them. It is trustworthy) of this given hypothesis.

| | Strongly disagree | Disagree | Neutral | Agree | Strongly agree | Unable to assess |
|---|---|---|---|---|---|---|
| The hypothesis is valid **scientifically** | ○ | ○ | ○ | ○ | ○ | ○ |
| The hypothesis is valid **clinically**, i.e., sound basis clinically | ○ | ○ | ○ | ○ | ○ | ○ |
| The hypothesis is **valid** (i.e., use this as an overall one instead of the above two subitems) | ○ | ○ | ○ | ○ | ○ | ○ |

*Q1.5.* The **potential benefits and risks** (Do the potential advantages to the potential stakeholders outweigh the costs and dangers?) for stakeholders, who will be the beneficiaries if this hypothesis can be translated into a large-scale study, will be evaluated by the following metrics.

| | Strongly disagree | Disagree | Neutral | Agree | Strongly agree | Unable to assess |
|---|---|---|---|---|---|---|
| The successful testing of this hypothesis will bring significant **benefits** to targeted audiences (e.g., patients, providers) | ○ | ○ | ○ | ○ | ○ | ○ |
| The testing of this hypothesis will bring no **risks** or tolerable risks to targeted audiences considering the benefits | ○ | ○ | ○ | ○ | ○ | ○ |
| The successful testing of this hypothesis will bring significant **benefits** to targeted audiences (e.g., patients, providers), i.e., the overall benefits outweigh the **risks** | ○ | ○ | ○ | ○ | ○ | ○ |

### Block 7

*Q2.1.* The following metrics will evaluate the **significance** (the quality of being important. The specific aspects that can be considered include medical needs, the future directions of the field, the target populations, costs, and benefits) of this hypothesis, i.e., the significance of a study used to test this hypothesis.

| | Strongly disagree | Disagree | Neutral | Agree | Strongly agree | Unable to assess |
|---|---|---|---|---|---|---|
| The hypothesis focuses on addressing **established medical needs**, e.g., a major medical problem affecting a relatively large population or the potential magnitude of improvement via testing this hypothesis for a severe condition | ○ | ○ | ○ | ○ | ○ | ○ |
| The test results of this hypothesis have the potential to impact the **future direction** of clinical practice positively | ○ | ○ | ○ | ○ | ○ | ○ |
| The test results of this hypothesis have the potential to impact the **target population** positively on average | ○ | ○ | ○ | ○ | ○ | ○ |
| The test of this hypothesis will be a worthwhile effort regarding the **cost** and benefit | ○ | ○ | ○ | ○ | ○ | ○ |
| Overall, this hypothesis is **significant** considering medical needs, cost and benefits, target population, and | ○ | ○ | ○ | ○ | ○ | ○ |

| | Strongly disagree | Disagree | Neutral | Agree | Strongly agree | Unable to assess |
|---|---|---|---|---|---|---|
| future directions of clinical practice. | | | | | | |

*Q2.5.* The following metrics will evaluate the **novelty** (the quality of being new and original) of a study to test this given hypothesis.

| | Strongly disagree | Disagree | Neutral | Agree | Strongly agree | Unable to assess |
|---|---|---|---|---|---|---|
| The test of a given hypothesis can lead to **innovation in medical practice** | ○ | ○ | ○ | ○ | ○ | ○ |
| The test of a given hypothesis can lead to **innovation methodology for clinical research** | ○ | ○ | ○ | ○ | ○ | ○ |
| The test of a given hypothesis can alter **previous findings**, i.e., has the potential to bring in paradigm shift in the field | ○ | ○ | ○ | ○ | ○ | ○ |
| The test of a given hypothesis can lead to **novel medical knowledge** | ○ | ○ | ○ | ○ | ○ | ○ |
| The test of a given hypothesis can lead to **new findings**, which can be incremental | ○ | ○ | ○ | ○ | ○ | ○ |
| Overall, this hypothesis is **novel** | ○ | ○ | ○ | ○ | ○ | ○ |

### Block 10

*Q3.1.* The **clinical relevance** (Is the hypothesis rooted within the clinical contexts? The specific aspects that can be considered include the potential impact on clinical practices, medical knowledge, and health policy) of a study to test a given hypothesis will be evaluated by the following metrics.

| | Strongly disagree | Disagree | Neutral | Agree | Strongly agree | Unable to assess |
|---|---|---|---|---|---|---|
| The test of a given hypothesis has the potential to impact **current clinical practice**, including patient safety, care quality | ○ | ○ | ○ | ○ | ○ | ○ |
| The test of a given hypothesis has the potential to impact our understanding of **medical knowledge** | ○ | ○ | ○ | ○ | ○ | ○ |
| The test of a given hypothesis has the potential to impact **health policy** | ○ | ○ | ○ | ○ | ○ | ○ |
| Overall, this hypothesis is **clinically relevant** | ○ | ○ | ○ | ○ | ○ | ○ |

*Q3.5.* The **feasibility** (How likely is the availability of resources [e.g., funds, eligible patients, etc.] needed to test the hypothesis ) of conducting a study to test a given hypothesis will be evaluated by the following metrics assuming the budget limit is 5 k US dollars and 0.5 years of a graduate student's time.

| | Strongly disagree | Disagree | Neutral | Agree | Strongly agree | Unable to assess |
|---|---|---|---|---|---|---|
| A study to test a given hypothesis is feasible regarding **needed cost**, i.e., needed resources or tools | ○ | ○ | ○ | ○ | ○ | ○ |
| A study to test a given hypothesis is feasible regarding **needed time** to conduct the study and to follow up | ○ | ○ | ○ | ○ | ○ | ○ |
| A study to test a given hypothesis is feasible regarding **scope**, i.e., a well-defined question | ○ | ○ | ○ | ○ | ○ | ○ |
| Overall, this hypothesis is **feasible** to test | ○ | ○ | ○ | ○ | ○ | ○ |

### Block 6

*Q4.1.* The **testability** (Given adequate resources [e.g., funds, eligible patients, etc.] can this hypothesis be tested) of a given hypothesis will be evaluated by the following metrics.

| | Strongly disagree | Disagree | Neutral | Agree | Strongly agree | Unable to assess |
|---|---|---|---|---|---|---|
| The hypothesis can be **tested** in an ideal setting | ○ | ○ | ○ | ○ | ○ | ○ |
| There are an adequate **number of patients** to choose from to participate in a | ○ | ○ | ○ | ○ | ○ | ○ |

| | Strongly disagree | Disagree | Neutral | Agree | Strongly agree | Unable to assess |
|---|---|---|---|---|---|---|
| study to test a given hypothesis | | | | | | |
| Overall, this hypothesis is **testable** | ○ | ○ | ○ | ○ | ○ | ○ |

*Q4.5.* The **clarity** (The quality of being coherent, transparent, and intelligible regarding the purposes, focused groups, variables, and their relationships within the hypothesis) of a given hypothesis will be evaluated by the following metrics.

| | Strongly disagree | Disagree | Neutral | Agree | Strongly agree | Unable to assess |
|---|---|---|---|---|---|---|
| The hypothesis provides clear **purpose**(s) | ○ | ○ | ○ | ○ | ○ | ○ |
| The hypothesis identifies **focused group(**s) | ○ | ○ | ○ | ○ | ○ | ○ |
| The hypothesis specifies **variable**(s) | ○ | ○ | ○ | ○ | ○ | ○ |
| The hypothesis specifies **the relationship**(s) between the variables under investigation | ○ | ○ | ○ | ○ | ○ | ○ |
| Overall, this hypothesis is **clear** | ○ | ○ | ○ | ○ | ○ | ○ |

### Block 9

*Q5.1.* The following metrics will evaluate the **ethicality** (Quality of being moral regarding the standards of right and wrong. One easy test is whether you trade the place with the potential participants if you are eligible) of a study to test this given hypothesis.

| | Yes | No | Unable to assess |
|---|---|---|---|
| There are no **ethical** concerns when conducting a study to test a given hypothesis, i.e., regarding patients, investigators, providers, and the conduction of the study | ○ | ○ | ○ |
| I will **trade my place** with a participant without hesitation in a study to test a given hypothesis | ○ | ○ | ○ |
| Overall, it is **ethical** to test this hypothesis | ○ | ○ | ○ |

*Q5.5.* The **interestingness** (Whether the hypothesis can catch the attention of peers, which will impact if the investigator can find potential collaborators for the project easily down the road) of this given hypothesis will be evaluated by the following metrics.

| | Strongly disagree | Disagree | Neutral | Agree | Strongly agree | Unable to assess |
|---|---|---|---|---|---|---|
| This hypothesis **interests** me | ○ | ○ | ○ | ○ | ○ | ○ |
| I will **pursue** the hypothesis if possible/feasible | ○ | ○ | ○ | ○ | ○ | ○ |

| | Strongly disagree | Disagree | Neutral | Agree | Strongly agree | Unable to assess |
|---|---|---|---|---|---|---|
| Overall, this is an **interesting** (i.e., the researcher should be able to find collaborators easily) hypothesis | ○ | ○ | ○ | ○ | ○ | ○ |

### Block 8

*Q6.1.* The overall quality score of the hypothesis on each dimension (1--the lowest; 5--the highest) will be evaluated by the following metrics:

| | 1 | 1.4 | 1.8 | 2.2 | 2.6 | 3 | 3.4 | 3.8 | 4.2 | 4.6 | 5 | Not Applicable |
|---|---|---|---|---|---|---|---|---|---|---|---|---|
| Validity | | | | | | | | | | | | ☐ |
| Significance | | | | | | | | | | | | ☐ |
| Novelty | | | | | | | | | | | | ☐ |
| Clinical relevance | | | | | | | | | | | | ☐ |
| Feasibility | | | | | | | | | | | | ☐ |
| Testability | | | | | | | | | | | | ☐ |
| Clarity | | | | | | | | | | | | ☐ |
| Ethicality | | | | | | | | | | | | ☐ |
| Potential benefits and risks | | | | | | | | | | | | ☐ |
| Interesting | | | | | | | | | | | | ☐ |
