## Supplementary material for "Data-driven hypothesis generation among inexperienced clinical researchers: A comparison of secondary data analyses with visualization (VIADS) and other tools": The coding principles (Supplemental Material 8)

**Time Coding Rules**

- Start Time: When data is pulled up and analysis begins
- End Time: After the researcher recaps the hypothesis and any further discussion about the hypothesis
- After one hypothesis ends, the next hypothesis begins either when the participant starts talking or begins moving their mouse and looking at the data. If nothing occurs for a long period of time, take half of the period.
- 1) if the duration differences between the two research assistants' results are < 10 seconds for any duration > 60 seconds, average the two measurements.
- 2) if the duration difference between the two research assistants' results > 10 seconds for any duration > 60 seconds and the difference is < 5% of the total duration, average the two measurements from the two research assistants.
    - If the difference is > 5%, check, revisit, discuss, and revise the final measurement.
- 3) if the duration difference is < 10 seconds for any duration < 60 seconds, and the difference is < 5% of the total duration, average the two measurements. ·
    - If the difference is > 5%, check, revisit, discuss, and revise the final measurement.
- 4) any other possibilities (the differences are larger than 5% of duration), please revisit, check, discuss, and revise accordingly.
- Averages are calculated by adding all the durations and then dividing by the number of hypotheses. (5 and above round up)
- Prep time is used for VIADS participants only and is used to measure the amount of time it takes from when they start looking into making a hypothesis to when the final graph is made before they make a hypothesis (as participants often use multiple graphs to gauge

what gives them the best ideas for a hypothesis). Start time for prep time is when the participant hits analyze on the data set for the first time. Data uploads during prep time are taken out of the duration of the prep time, as they do not contribute to hypothesis generation.

- If the researcher asks, "Would you like to continue" and the participant wants to, then that start time will begin when the researcher first speaks.
- If there is a data upload in between hypotheses, the time before data upload and right after the participant hits analyze, up until the last graph is made before the next hypothesis is added together to make another separate prep time.
- If hypotheses are made in rapid succession, once the recap of each is made, they are averaged.
- Tool learning in the beginning before the participants begin to analyze the data is not included in the time, as participants in VIADS often need time to get used to the program.
